# Selective prediction as a triage gate for primary-care depression screening: quantifying and mitigating selection bias in CHARLS-2011

**DOI:** 10.64898/2026.07.17.26357845

**Authors:** Zijian Wang, Yaqing Liu

**Affiliations:** School of Medicine and Health Management, Tongji Medical College, Huazhong University of Science and Technology, Wuhan, Hubei 430030, China

**Keywords:** selective prediction, selection bias, CHARLS, depression, predictor–selector decoupling, uncertainty quantification, classification and regression tree, triage, clinical decision support, health management

## Abstract

**Background:** Primary care in China lacks structured mental-health assessment, and the machine-learning models that could support such screening are typically developed on heavily selected samples. Cumulative inclusion and exclusion criteria, though usually treated as neutral data-cleaning steps, can create heterogeneity in predictive reliability among retained participants. Using the China Health and Retirement Longitudinal Study (CHARLS) 2011 baseline, we quantified how selection funnels distort epidemiological associations and inflate machine-learning metrics, and tested selective prediction as mitigation.

**Methods:** Using the CHARLS 2011 baseline with temporal external validation in CHARLS-2018, we built a four-level selection funnel (L0–L3), evaluated five classifiers with nested cross-validation and SMOTE, and compared model-embedded uncertainty with a decoupled predictor–selector framework; XGBoost cross-validation residuals drove risk stratification and classification and regression tree (CART) rules.

**Results:** Sample sizes fell from L0 n=17,705 to L3 n=4,256 (24.0%). The cancer–depression odds ratio attenuated from 1.78 (95% CI 1.32–2.41) to 1.39 (0.74–2.63), losing significance. AUC rose with selection but not after multiple-comparison correction, whereas calibration error increased for four of five models. Model-embedded uncertainty succeeded only for XGBoost; with the decoupled XGBoost residual selector, all five models achieved selective prediction at approximately 20% coverage (test AUC 0.90, 95% CI 0.85–0.95), abstaining on approximately 80% of cases for individual safety. Risk stratification was stable (residual Spearman correlations >0.95; multi-seed Jaccard 0.88), and CART rules used self-rated health, education, pain, and marital status.

**Conclusions:** The findings support a deployable primary-care triage pathway: a four-variable rule identifies patients suitable for algorithm-assisted scoring (approximately 20% coverage) and routes the remainder to human evaluation. Methodologically, cumulative selection bias produces a dual distortion: epidemiological associations are compressed and machine-learning metrics inflated. Selective prediction is limited mainly by uncertainty-indicator design. Performance metrics should be reported with selection level, coverage, and calibration trajectory. Decoupled selective prediction with CART rule extraction provides an actionable framework for quality-controlled, tiered-care deployment.

## Introduction

In China, primary care institutions are responsible for chronic disease management and initial mental health screening for older adults, yet general practitioners often receive limited psychiatric training and specialist referral resources are scarce [31]. The current national basic public health service package provides free annual physical examinations for adults aged 65 years and older, but these visits focus primarily on somatic indicators and rarely include structured mental health assessment [32]. Against this backdrop, a decision-support tool that uses only a few routinely collected variables and explicitly flags which patients are suitable for algorithm-assisted scoring and which require human evaluation is likely to be more useful in grassroots settings than a black-box model that issues probabilities for everyone.

Conceptually, this tool is best understood as a lightweight clinical decision-support node embedded within chronic-disease management pathways; it is not designed as a standalone screening test [33,34].

### Machine-learning depression prediction in CHARLS: promise and hidden bias

Machine-learning models for late-life depression in CHARLS achieve moderate discrimination, but most carry a high risk of bias, especially around participant selection [3–6].

### Selection criteria and their hidden cost

Inclusion and exclusion criteria are standard epidemiological tools intended to ensure data completeness and model feasibility. Investigators usually restrict analyses to participants with complete outcomes, valid exposures, and no disqualifying comorbidities. Yet a funnel may begin with more than 17,000 participants and end with fewer than 2,000 after sequential exclusions [4]. Authors often report the number excluded at each stage but rarely compare the epidemiological profile of retained participants with that of the excluded majority. They also rarely ask whether post-selection associations differ from unselected associations. This omission is problematic. Conditioning on variables that are causes of both exposure and outcome induces collider stratification, which can create or distort associations [7]. When applied to depression prediction, excluding participants with severe cognitive impairment may reshape the apparent importance of education, marital status, or chronic disease burden. Those excluded may also have higher depression prevalence. The central question, then, is whether reported associations and metrics from a highly selected subset are generalizable to the original target population.

### Three overlooked assumptions

Machine-learning depression prediction rests on three rarely examined assumptions that have profound validity implications.

Assumption 1: selection only restricts range without changing association direction. Every selection decision conditions the data and reshapes the covariance structure of what remains. Collider bias can induce spurious associations and materially change effect estimates [7]. For clinical deployment, this means that an association estimated in a selected analytic sample may not describe the patients a screening tool will actually encounter.

Assumption 2: reported performance metrics, especially AUC, are objective reflections of model ability. CHARLS-based studies commonly report AUCs of 0.71–0.80 [3,5]. Yet AUC is computed on the selected sample. It does not reflect the difficulty of excluded cases or performance on the original cohort. In imbalanced datasets, AUC can produce misleadingly optimistic estimates when combined with synthetic oversampling and no external validation [8]. A high AUC reported from a selected funnel is therefore not, by itself, evidence that a model is ready for clinical use.

Assumption 3: most models are equally suited to selected data. Different algorithms encode different inductive biases that may interact differently with selection-induced distribution shifts. A model that performs well on a homogeneous selected subpopulation may fail on the heterogeneous source population [6]. The literature largely ignores this model–data interaction and treats evaluation as model agnostic, ignoring the specific conditions under which each model–data pair operates. This assumption is especially problematic given documented high risk of bias: 73% of 81 reviewed studies were rated high risk, with inadequate missing-data handling and nontransparent participant selection as leading domains [6,38]. Deployment therefore requires knowing for whom a model can be trusted, not only how well it performs on average.

### Selective prediction as a boundary-defining tool

Selective prediction, or classification with a reject option, allows a classifier to abstain when its confidence falls below a threshold [9,10,37]. The theoretical promise is that rejecting uncertain predictions can guarantee higher accuracy on the retained subset. Whether this promise holds in epidemiological selection pipelines depends on whether the uncertainty indicator truly captures predictive reliability, an issue that has not been examined in this context. In model-embedded approaches, the same algorithm that produces predictions also produces uncertainty scores, risking semantic coupling: if model confidence is uncalibrated, the uncertainty score is contaminated by the same distortion. This study asks a further question: when model-embedded uncertainty indicators fail, can a decoupled predictor–selector framework rescue selective prediction by having an independent model assess predictive reliability?

### Translational objectives

Against this backdrop, this study follows a translational path with three nested objectives.

Objective 1: detect the bias. Quantify how estimated associations between chronic diseases and depression evolve across selection levels, and assess whether machine-learning performance metrics are conditional on the selection funnel. We use monotonic-trend analysis to formalize the relationship between selection and performance. Specifically, we build the four-level L0–L3 funnel in CHARLS 2011, examine how each level reshapes logistic-regression odds ratios, and evaluate five machine-learning classifiers. Page’s L test formally tests whether AUC, Brier score, and expected calibration error change systematically with selection strength.

Objective 2: mitigate the bias. Reveal whether selective prediction effectiveness is driven primarily by uncertainty-indicator design. We compare model-embedded uncertainty with a decoupled predictor–selector framework containing nine candidate selectors. We then use XGBoost cross-validation residuals to stratify the full L3 sample into domains of reliable and unreliable prediction.

Objective 3: enable clinical deployment. Convert black-box rejection into interpretable white-box rules that answer the clinical question of whom the model is truly fit to predict. A classification and regression tree extracts split rules and characterizes risk groups. Multi-parameter and multi-seed stability checks ensure that rules are not artifacts of implementation choices. The final pathway moves from patient triage to stratified intervention: classify patients with four easily collected clinical variables, apply the screened model to score depression risk in the reliable-prediction domain, and trigger human evaluation in the high-uncertainty domain.

## Methods

### Study design and data source

This study used the China Health and Retirement Longitudinal Study (CHARLS), a nationally representative longitudinal survey of Chinese adults aged 45 years and older [2]. Baseline data were collected between June 2011 and March 2012 across 150 county-level units in 28 provinces. We restricted analyses to the 2011 baseline survey so that cumulative selection effects could be isolated from attrition-related confounding introduced in later waves. The CHARLS protocol was approved by the Peking University Institutional Review Board (IRB00001052–11015), and all participants provided written informed consent [2]. We accessed the publicly available CHARLS-2011 baseline dataset through the official data portal [36]. Reporting follows the Strengthening the Reporting of Observational Studies in Epidemiology (STROBE) guidelines for cross-sectional studies.

### Four-level selection funnel (L0–L3)

We formalized the exclusion process common to almost every observational analysis by constructing a four-level selection funnel [7]. L0 contained the largest complete sample after missing data were addressed with multiple imputation by chained equations (MICE) [11,39]. Missing-data patterns are visualized in Supplementary Figure S2. Predictive mean matching was used for continuous variables, logistic regression for binary variables, and multinomial regression for categorical variables. We generated five imputed datasets and combined estimates using Rubin’s rules. For the machine-learning pipeline, a single imputed realization was retained for computational tractability. Consequently, L0 machine-learning results are conditional on that realization and are not directly comparable with the complete-case L1–L3 levels; the sensitivity analysis only shows that results were insensitive to the choice among imputations, not that L0 is a true population reference. L0 therefore serves as a reference population, not a truly complete cohort; some records had item-missing patterns too extensive for reliable imputation. L1 retained participants aged 60 years or older. L2 further required complete CES-D 10-item responses so that the depression score could be computed. L3 additionally required complete values for the key sociodemographic covariates education, health insurance status, and residence. Cumulativeness is essential: each level compounds the selection effect of the previous level. Supplementary Table S1 shows sample sizes and demographic characteristics by level, and Supplementary Figure S1 provides a flowchart of the screening funnel. No formal sample-size calculation was performed because this study is a secondary analysis of the existing CHARLS 2011 cohort; the L3 analytic sample (n=4,256; approximately 1,770 depression events) provides more than 50 events per predictor, exceeding common empirical thresholds for binary prediction-model development.

### Association analysis: testing Assumption 1

The outcome was depressive symptoms, operationalized with the Center for Epidemiologic Studies Depression scale (CES-D) [12,13]. The scale is a 10-item short-form self-report instrument with high internal consistency in Chinese populations; Cronbach’s α typically exceeds 0.85. Following established CHARLS practice, we used a cutoff of ≥10 to define clinically significant depressive symptoms [1,14]. Exposure variables were five chronic-disease indicators: cancer, chronic lung disease, hypertension, diabetes/high blood sugar, and chronic disease count (variable definitions are provided in Supplementary Table S5). The individual conditions were based on respondent-reported physician diagnoses and coded as binary variables (0/1) [15].

We fitted a separate logistic regression model for each exposure. The binary depression indicator was the outcome. Covariates were the exposure, age (continuous), sex (female as reference), and education (categorical). Model specification was identical across selection levels (L0–L3). Observed differences in association estimates therefore reflect changes in sample composition under a fixed model form. From each model we extracted adjusted odds ratios (ORs) and 95% confidence intervals (CIs) using the Wald method. We used Cochran’s Q to test heterogeneity of ORs across levels under the null hypothesis of constant ORs.

### Machine-learning evaluation: testing Assumption 2

We evaluated five models: logistic regression (LR), random forest (RF), XGBoost (XGB), LightGBM (LGB), and gradient boosting (GB). LR served as an interpretable linear baseline [16]. RF reduced overfitting through bootstrap aggregation [17]. XGB implemented regularized gradient boosting [18]. LGB used histogram-based leaf-wise boosting with lower computational cost [19]. GB provided a classical boosting reference.

The feature set included all variables in the logistic-regression specification plus marital status, residence, self-rated health, activities of daily living (ADL) limitations, instrumental activities of daily living (IADL) limitations, smoking status, and alcohol use. Categorical variables were one-hot encoded and continuous variables standardized. No feature selection was performed; each model’s built-in regularization managed dimensionality. This design ensured that machine-learning models had access to the same information as the logistic-regression model, enabling fair comparison of how different algorithms used the same feature space under selection-induced distribution shift. Avoiding feature selection also prevented an additional source of bias that could confound comparisons across selection levels.

Within each level, 15% of observations were set aside as an independent test set and were not used for tuning. The remaining 85% were used for grid-search hyperparameter tuning with 5-fold stratified cross-validation, scored by AUC [20]. Synthetic Minority Over-sampling Technique (SMOTE) [21] was embedded in the imbalanced-learn pipeline and applied only to the training fold of each cross-validation split; validation and test sets retained their original distributions. The entire procedure was repeated across seven random seeds, producing repeated-measure data for subsequent trend tests. Monotonic trend analysis used Page’s L test with seed as block and selection level as ordered treatment, which is designed for repeated-measures ordered designs. Because AUC, Brier score, and ECE [22,23] are bounded metrics and the number of seeds is modest, the normal-approximation p-values are treated as approximate; significance decisions rely on the multiple-comparison-adjusted results.

AUC was the primary metric of model discrimination. It measures the probability that a randomly chosen case is ranked above a randomly chosen non-case and is independent of outcome prevalence. Although this prevalence independence protects AUC from simple class-ratio changes, AUC remains sensitive to changes in sample homogeneity; as selection removes harder-to-separate cases, AUC can rise mechanically. We focused on the monotonic trend in AUC to verify the conditional relationship between selection strength and discrimination.

### Selective prediction framework: testing Assumption 3

We implemented two frameworks. Clinically, this amounts to giving the model a right to abstain: it issues scores only for patients whose predictions are demonstrably reliable.

#### Model-embedded uncertainty

In the model-embedded uncertainty analysis, each algorithm used its own uncertainty indicator. For XGBoost, we designed a custom indicator based on cross-validated quantile residual intervals. Nested 3-fold stratified cross-validation was performed within the training set. In each fold, an XGBoost classifier was trained on the other two folds and produced out-of-fold predicted probabilities. These out-of-fold probabilities were concatenated across folds to cover all training samples. Absolute out-of-fold residuals were computed, and two XGBoost quantile regressors (α=0.1 and α=0.9) were trained to predict the 10th and 90th percentiles of residuals. The test-set uncertainty score was defined as the width of the quantile interval (q90 − q10). This design ensured that uncertainty assessment did not rely on in-sample predictions, which would systematically underestimate residuals because of information leakage. Each sample’s residual came from a model trained without that sample. For LR we used predictive entropy. RF used tree-to-tree probability variance. LGB and GB used 1 − max(probability).

We constructed rejection–coverage (RC) curves by varying the rejection threshold on the validation set. Coverage ranged from 100% to 17.5% in 2.5% steps. For each model, we identified the coverage that maximized AUC, ensuring that comparisons were not biased by subjective target selection. Each point on an RC curve represents a hypothetical scenario in which the model predicts only on the X% of samples it deems most reliable.

#### Decoupled predictor–selector

In the decoupled predictor–selector framework, the predictor (classification model) and selector (uncertainty quantifier) are largely decoupled. We evaluated nine candidate selectors, each producing an uncertainty score independent of the predictor’s internal state: XGB residual regression, XGB correctness classification, ensemble disagreement, k-nearest-neighbor local density, predictor self-confidence, kernel density estimation, Mahalanobis distance, autoencoder reconstruction error, and Gaussian mixture model density. Subsequent analyses used XGBoost cross-validation residuals as the external selector. For each predictor (LR, RF, XGB, LGB, GB), the nine selectors were evaluated in stratified 5-fold cross-validation. The selector-selection logic included a smoothness preference: if a smooth selector (such as Gaussian mixture model density or the XGBoost cross-validation residual interval) yielded AUC within 0.005 of the best, it was preferred. This encouraged clinically interpretable, low-variance uncertainty estimates. The final selected selector was applied to the held-out test set. Test-set RC curves were computed at 2.5% resolution. All supervised selectors (XGB residual regression, XGB correctness classification) required cross-validated signals to avoid information leakage. Predictors were trained on two inner folds and evaluated on the third, yielding cross-validation residuals or correctness labels for the entire training set. Selectors were then trained on these signals and never saw the same data on which the predictor was trained.

#### Robust-residual risk stratification and CART interpretable rules

To answer the clinical question of which samples the model is truly fit to predict, we first computed out-of-fold residuals from XGBoost under five distinct hyperparameter configurations. Each configuration used nested 5-fold × 3-fold cross-validation to prevent information leakage. The median residual across configurations was taken as the robust residual. Lower residuals indicated more reliable predictions; higher residuals indicated more confused predictions.

Risk groups were defined directly from ranks of the raw robust residuals. The sample was split into low-, medium-, and high-risk tertiles (approximately one-third each) at the 33rd and 67th percentiles. Classification and regression tree (CART) was used only for rule extraction and phenotyping, fitting a regression tree (max_leaf_nodes=8, min_samples_leaf=75) with robust residual as the outcome and 15 clinical features as predictors. Rules for each branch were extracted with scikit-learn.tree.export_text to characterize the clinical profile of each risk group. CART was not used for extrapolation, and risk grouping did not depend on CART-predicted residuals.

Validation used independent-test-set logistic-regression AUC. Specifically, low, medium, and high groups were defined within the training set by robust-residual rank; separate logistic-regression classifiers were trained in each group; and AUC was evaluated in the corresponding groups of the independent test set. Because groups were defined by residual rank, these within-group AUCs characterize separability conditional on the grouping and should not be read as independent evidence of model performance; the near-perfect and near-random values are expected artifacts of the residual-based split. Finally, we performed multi-parameter (five hyperparameter configurations) and multi-seed (ten random seeds) stability checks: Spearman correlations of residuals across configurations and Jaccard similarity of high-risk group sample sets. If correlations exceeded 0.90 and Jaccard similarities exceeded 0.90, the risk stratification could be regarded as a stable property of the data across implementations.

### Multiple comparison correction

Given the large number of statistical tests (5 exposures × 4 levels = 20 OR estimates; 5 models × 4 metrics = 20 Page’s L tests; 9 selectors × 5 predictors = 45 selector evaluations in the decoupled framework), we applied multiple-comparison correction. For the primary confirmatory tests (OR trend and AUC monotonic-trend tests), we used Bonferroni correction, dividing the nominal α of 0.05 by the number of tests within each family. For exploratory analyses, including selector comparison and sensitivity analyses with alternative CES-D cutoffs, we used the Benjamini–Hochberg false discovery rate (FDR) procedure at q=0.10 [24]. This dual approach balances Type I error control and statistical power and reflects the distinction between hypothesis testing and hypothesis generation in epidemiological research.

### External validation

To evaluate the temporal stability of the CART-based triage workflow, we applied the four-variable CART rules derived from CHARLS-2011 to the harmonized CHARLS-2018 sample. CHARLS-2018 is the fourth wave of the same cohort and is affected by attrition and changes in chronic-disease measurement; therefore, this validation focused on whether the CART low-risk group retained acceptable discrimination in the 2018 sample. Group-specific AUCs were computed with XGBoost, and group-level AUCs remained stable across multiple seeds.

## Results

### Funnel characteristics

Applying the predefined inclusion and exclusion criteria across four levels (L0–L3) produced substantial sample attrition. As shown in Supplementary Table S1, L0 contained 17,705 participants after MICE imputation. L1 retained 7,669 (43.3% of L0); L2 retained 6,594 (37.2%); and L3 retained 4,256 (24.0%). Mean age increased from 59.1 years at L0 to 68.6 years at L3. The proportion of female participants rose from 47.8% to 53.9%. Depression prevalence rose from 36.9% at L0 to 41.6% at L3, accompanied by large shifts in family-support score (SMD = −0.638, large effect), marital status (SMD = 0.474, moderate effect), and number of chronic conditions (SMD = −0.190, small effect). These compositional changes altered the joint distribution of exposures, outcome, and confounders, directly affecting epidemiological effect estimates and predictive model performance.

### Association distortion (Assumption 1)

Logistic-regression models fitted separately for each of the five chronic-disease indicators at each selection level revealed differential association compression. Table 1 presents odds ratios and 95% confidence intervals; full results for all exposures are provided in Supplementary Table S2, and the corresponding odds-ratio drift heatmap is shown in Supplementary Figure S3.

**Table 1.** Association analysis. Odds ratios (95% CI) for depression across selection levels L0–L3.

| Exposure | L0 OR (95% CI) | L1 OR (95% CI) | L2 OR (95% CI) | L3 OR (95% CI) |
| --- | --- | --- | --- | --- |
| Cancer | 1.781 [1.317–2.410] | 1.315 [0.799–2.164] | 1.371 [0.816–2.301] | 1.393 [0.737–2.632] |
| Chronic disease count | 1.450 [1.416–1.486] | 1.416 [1.367–1.468] | 1.412 [1.361–1.464] | 1.422 [1.358–1.489] |
| Chronic lung disease | 1.998 [1.804–2.214] | 1.749 [1.519–2.013] | 1.754 [1.518–2.027] | 1.883 [1.575–2.251] |
| Diabetes/high blood sugar | 1.414 [1.237–1.616] | 1.424 [1.176–1.723] | 1.395 [1.147–1.696] | 1.433 [1.119–1.834] |
| Hypertension | 1.255 [1.165–1.350] | 1.195 [1.075–1.327] | 1.180 [1.059–1.315] | 1.214 [1.062–1.388] |

### Machine-learning performance across models and selection levels

The five machine-learning models (LR, RF, XGB, LGB, GB) were trained and evaluated at each selection level (L0–L3). Table 2 presents the full metric matrix; complete metrics across all levels are provided in Supplementary Table S3.

**Table 2.**
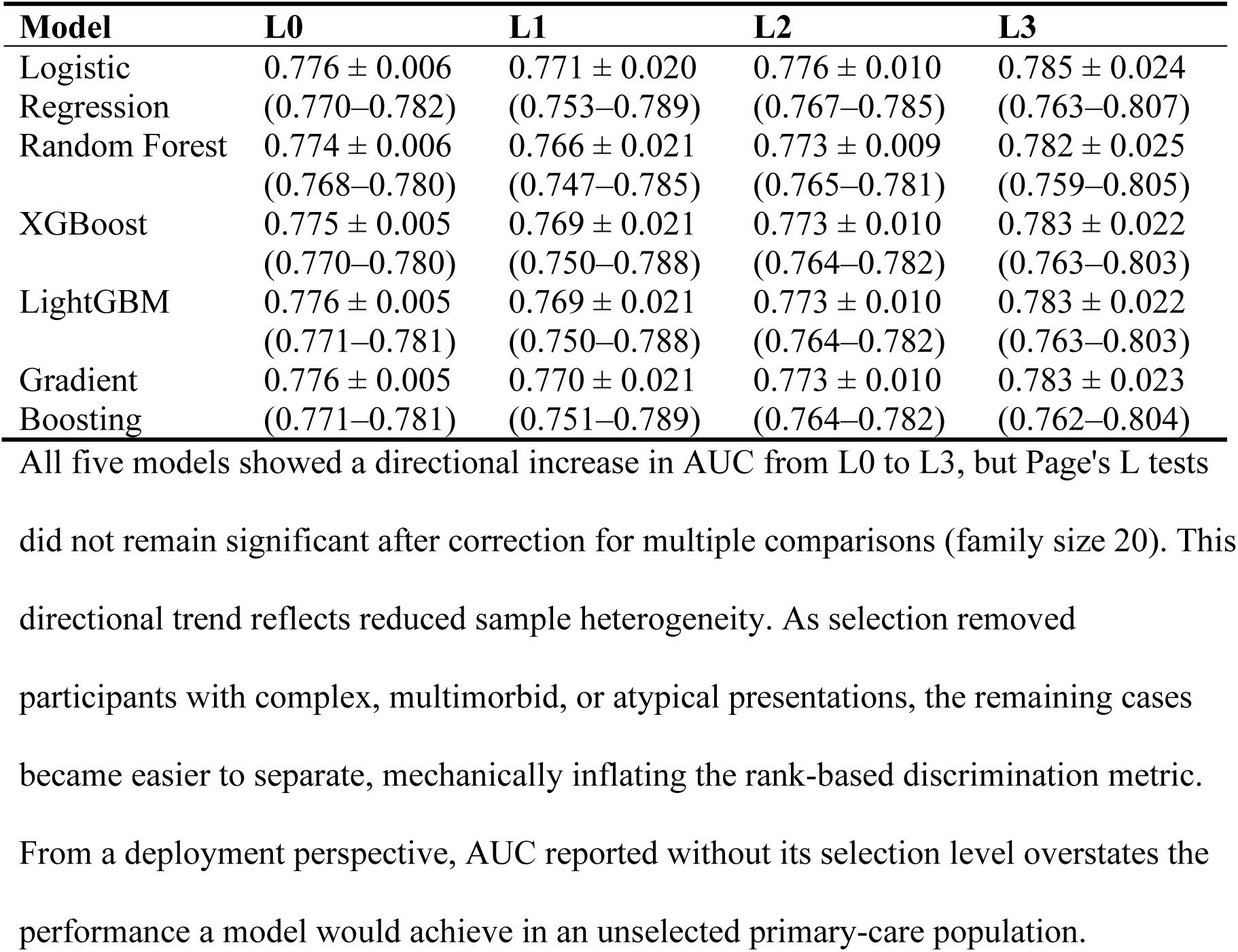
Test-set AUC (mean ± standard deviation across seven random seeds) by model and selection level.

### Selective prediction

#### Model-embedded uncertainty

Table 3 presents key operating points from the model-embedded selective-prediction analysis. We did not prespecify a fixed AUC target. Instead, we identified the coverage that maximized AUC in cross-validation and validated the performance at that coverage on the test set.

**Table 3.** Selective prediction. Key operating points on rejection–coverage curves.

| Model | CV coverage | CV AUC | Test coverage | Test AUC | Precision | Recall | Accuracy |
| --- | --- | --- | --- | --- | --- | --- | --- |
| Logistic Regression | 100.0% | 0.771 | 100.0% | 0.791 | 0.682 | 0.676 | 0.734 |
| Random Forest | 100.0% | 0.771 | 95.0% | 0.793 | 0.706 | 0.648 | 0.741 |
| XGBoost | 32.4% | 0.848 | 52.5% | 0.831 | 0.771 | 0.614 | 0.776 |
| LightGBM | 100.0% | 0.769 | 100.0% | 0.789 | 0.714 | 0.625 | 0.739 |
| Gradient Boosting | 100.0% | 0.769 | 100.0% | 0.791 | 0.706 | 0.637 | 0.738 |

For logistic regression, random forest, LightGBM, and gradient boosting, the embedded uncertainty indicators, i.e., predictive entropy (LR), tree-to-tree variance (RF), and 1 − max(probability) (LGB, GB), failed to identify an AUC peak better than full coverage in cross-validation (optimal coverage = NaN). These generic uncertainty indicators could not effectively distinguish easy from difficult cases. On the test set, RF showed a small AUC gain at 95% coverage (0.7930). The other three models remained at 100% coverage, with no selective gain. This result suggests that selective prediction effectiveness depends on the specific pairing of uncertainty indicator and model architecture.

The RC curves explain the four failures. For XGBoost, the curve rose steeply at first: AUC increased rapidly as coverage fell from 100% to about 50%, then gains decelerated. This steep-then-flat shape is the signature of an informative uncertainty indicator. The model quickly identified highly uncertain cases and abstained on them; remaining predictions came from a well-characterized domain. For LR, RF, LGB, and GB, the curves were flat or monotonic, with no discernible inflection point where selective abstention produced meaningful AUC improvement. The absence of an inflection point indicates that their uncertainty indicators failed to rank samples by prediction difficulty. The structural gap between the XGBoost curve and the four flat curves is the central visual evidence of this analysis. It suggests that effective selective prediction is a property of the specific pairing of uncertainty indicator, model architecture, and data distribution, and may not be a generic property of machine-learning classifiers.

#### Decoupled predictor–selector

The picture changed markedly when an independent selector was introduced. Table 4 presents the results. A CV coverage of 0.200 was the threshold determined by optimization in cross-validation for the XGBoost residual selector. The high consistency of coverage across the five models indicates that XGBoost residuals, when used as an external selector, identified reliable samples with cross-model stability.

**Table 4.** Selective prediction. Decoupled predictor–selector results.

| Predictor | CV coverage | CV AUC | Test coverage | Test AUC | Precision | Recall | Accuracy | Test AUC 95% CI |
| --- | --- | --- | --- | --- | --- | --- | --- | --- |
| Logistic Regression | 19.9% | 0.893 | 20.0% | 0.901 | 0.86 | 0.892 | 0.876 | 0.85–0.95 |
| Random Forest | 19.9% | 0.893 | 20.0% | 0.895 | 0.852 | 0.904 | 0.876 | 0.84–0.94 |
| XGBoost | 19.9% | 0.891 | 20.0% | 0.888 | 0.851 | 0.892 | 0.871 | 0.83–0.94 |
| LightGBM | 19.9% | 0.89 | 20.0% | 0.894 | 0.851 | 0.892 | 0.871 | 0.84–0.94 |
| Gradient Boosting | 19.9% | 0.89 | 20.0% | 0.892 | 0.852 | 0.904 | 0.876 | 0.84–0.94 |

All five models paired with the XGBoost cross-validation residual quantile interval external selector achieved clear selective-prediction gains. Coverage was highly consistent across models (approximately 20.0%), CV AUC ranged from 0.8898 (GB) to 0.8934 (LR), and test AUC ranged from 0.8876 (XGB) to 0.9008 (LR). Test precision was approximately 0.85–0.86, recall approximately 0.89–0.90, and accuracy approximately 0.87–0.88. The approximately 20% operating point reflects a conservative, patient-safety-first strategy: predictions are returned only when the selector identifies a reliable domain, and uncertain cases are channeled to human evaluation, avoiding potentially misleading automated labels.

The methodological implication is that XGBoost cross-validation residuals are useful across model architectures, including XGBoost, LR, RF, LGB, and GB. When XGBoost cross-validation residuals were used as the external selector, the optimal coverage was highly consistent across models (approximately 20%), and about 80% of samples were consistently classified as high uncertainty. XGBoost cross-validation residuals therefore outperformed candidate selectors based on geometric density or model consensus.

This convergence (approximately 20% in both cross-validation and test sets) is strong evidence for the robustness of the decoupled selective-prediction framework. Different model architectures agreed on which samples were reliable; they differed only in how those samples were classified. This has clinical implications: in resource-constrained screening programs, one could first use XGBoost to identify approximately 20% of samples as reliable predictions and then apply a simple model such as LR for depression risk scoring, achieving an AUC of approximately 0.89 and precision of approximately 0.86.

**Figure 1.**
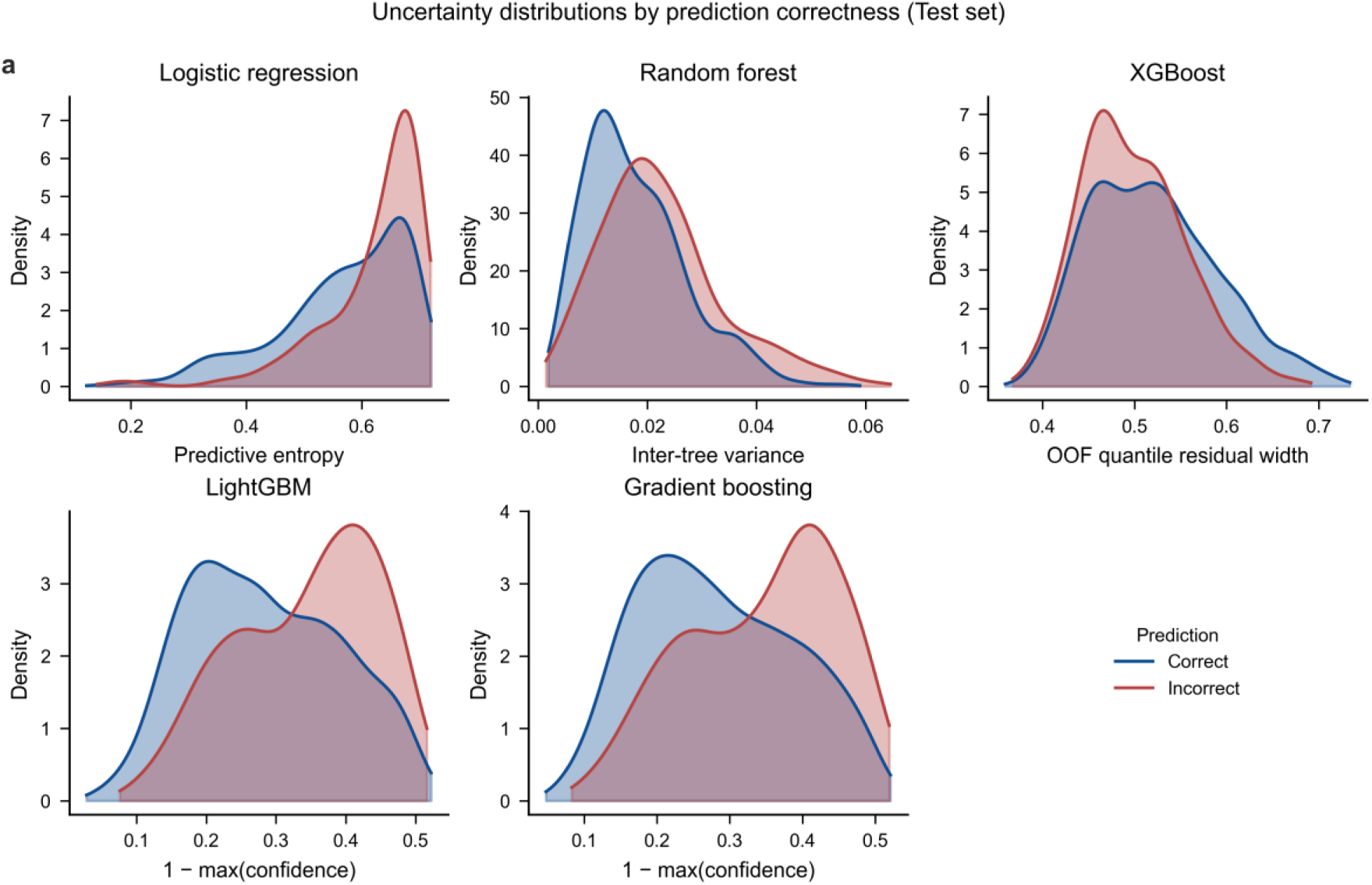
Distribution of model-embedded uncertainty scores for correct and incorrect predictions across five classifiers.

**Figure 2.**
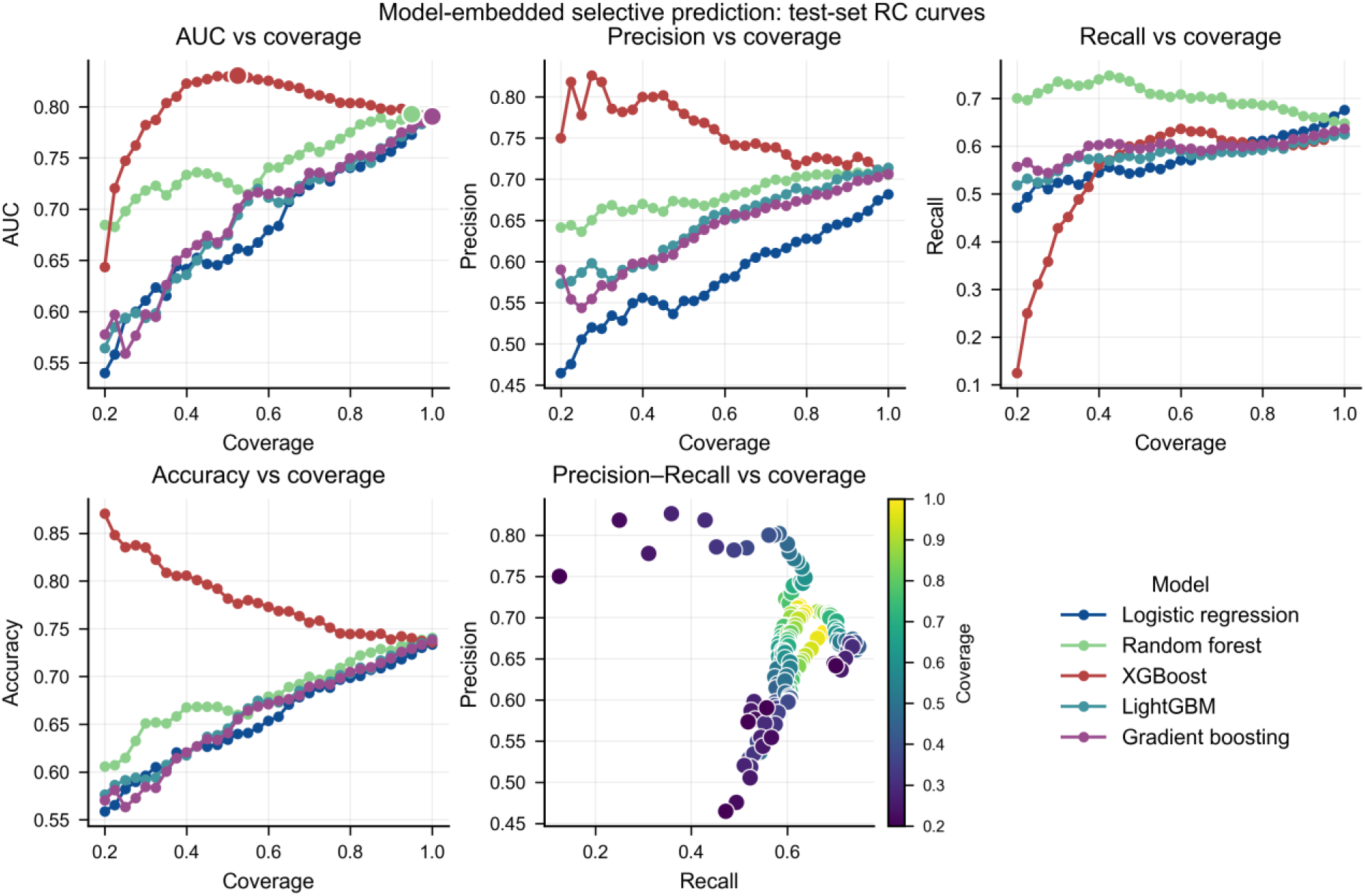
Model-embedded selective prediction: rejection–coverage curves for five classifiers.

**Figure 3.**
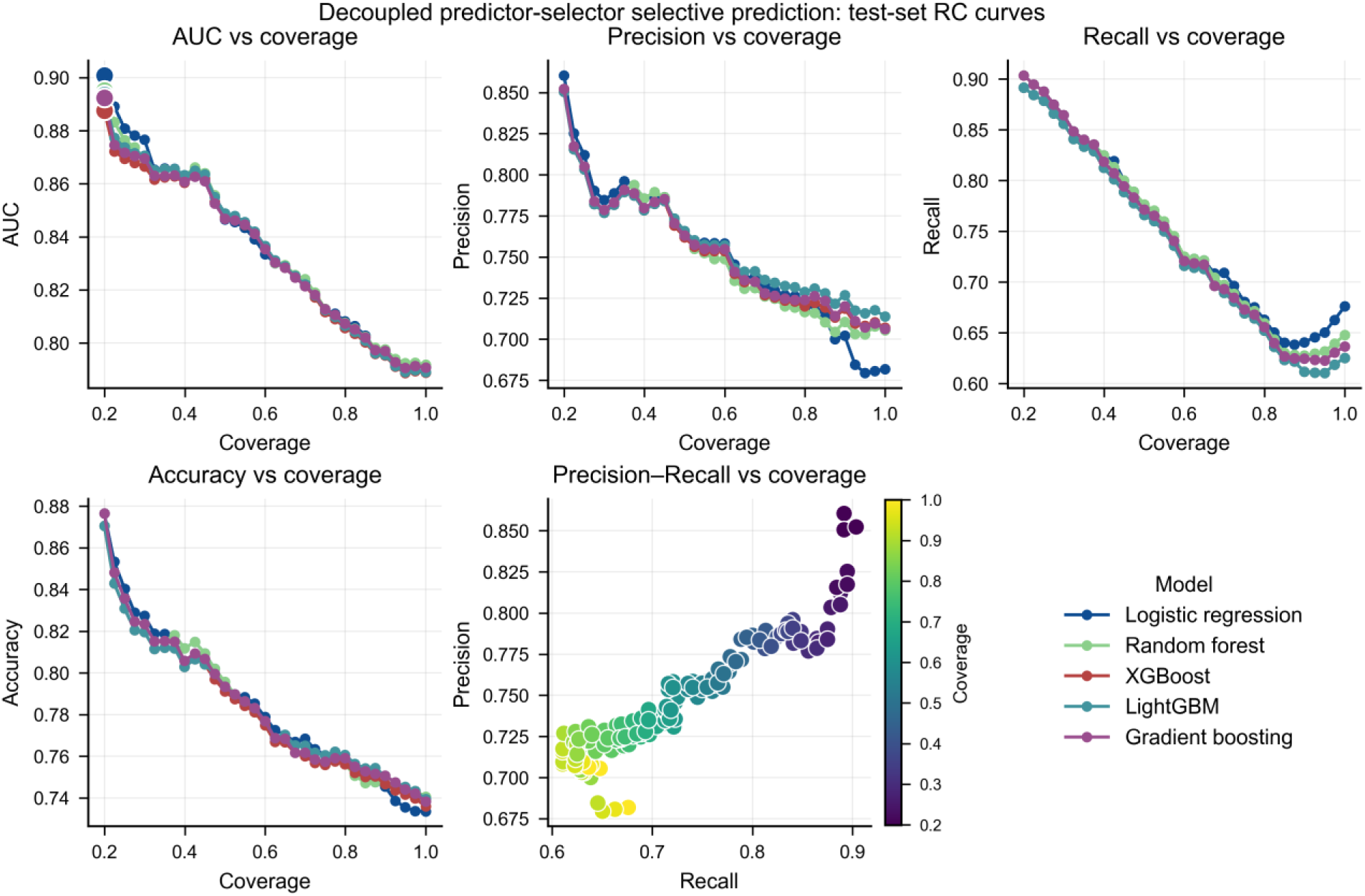
Decoupled predictor–selector selective prediction: test-set rejection– coverage curves using XGBoost cross-validation residuals as the external selector.

### Risk stratification and CART interpretable rules

#### Characteristic profiles of the three risk groups

Based on ranks of raw robust residuals, the L3 sample was divided into low-, medium-, and high-risk tertiles. A CART regression tree (max_leaf_nodes=8, min_samples_leaf=75) was fitted to extract interpretable split rules and characterize clinical phenotypes. Table 5 presents demographic and clinical characteristics by group.

**Table 5.**
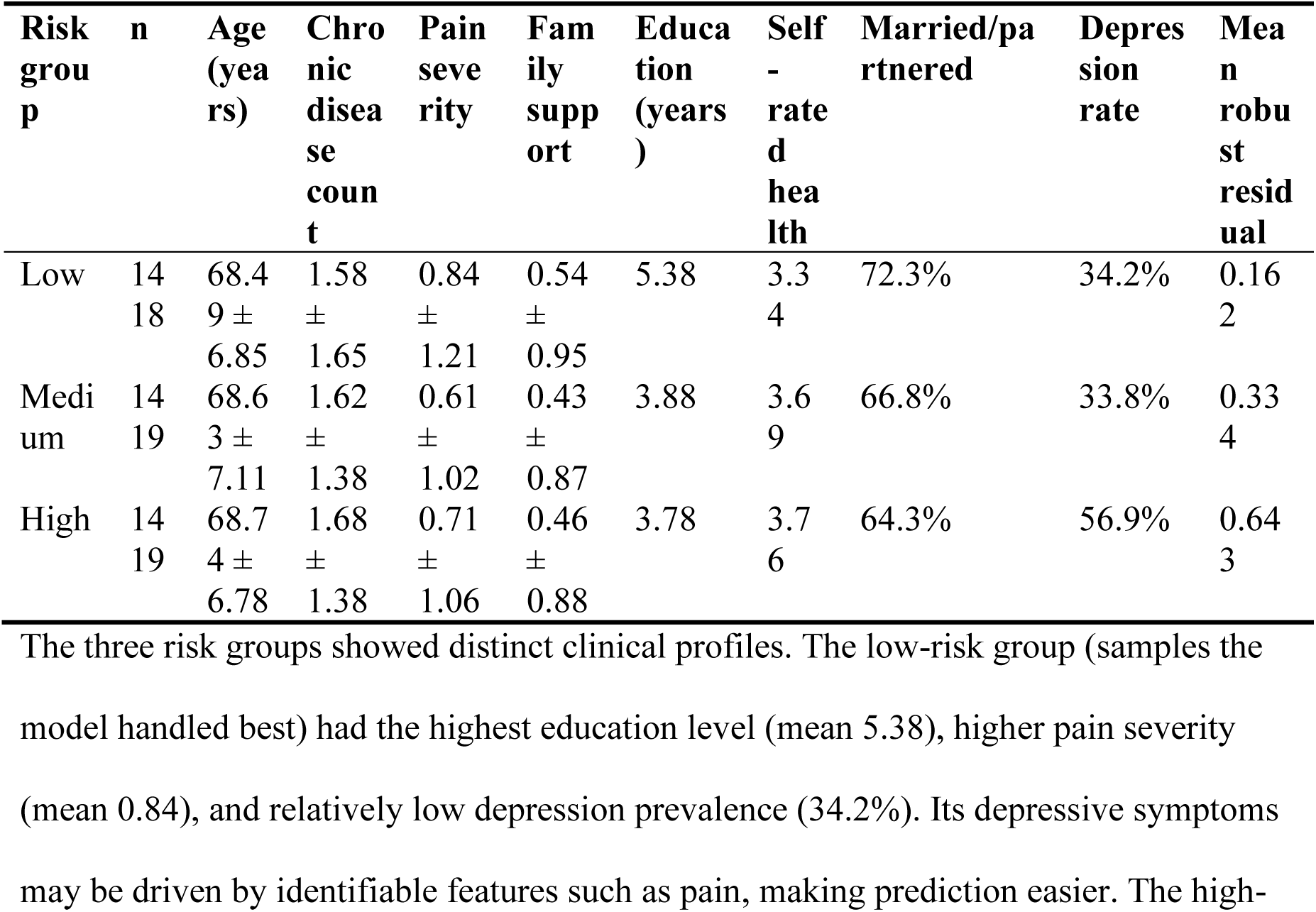
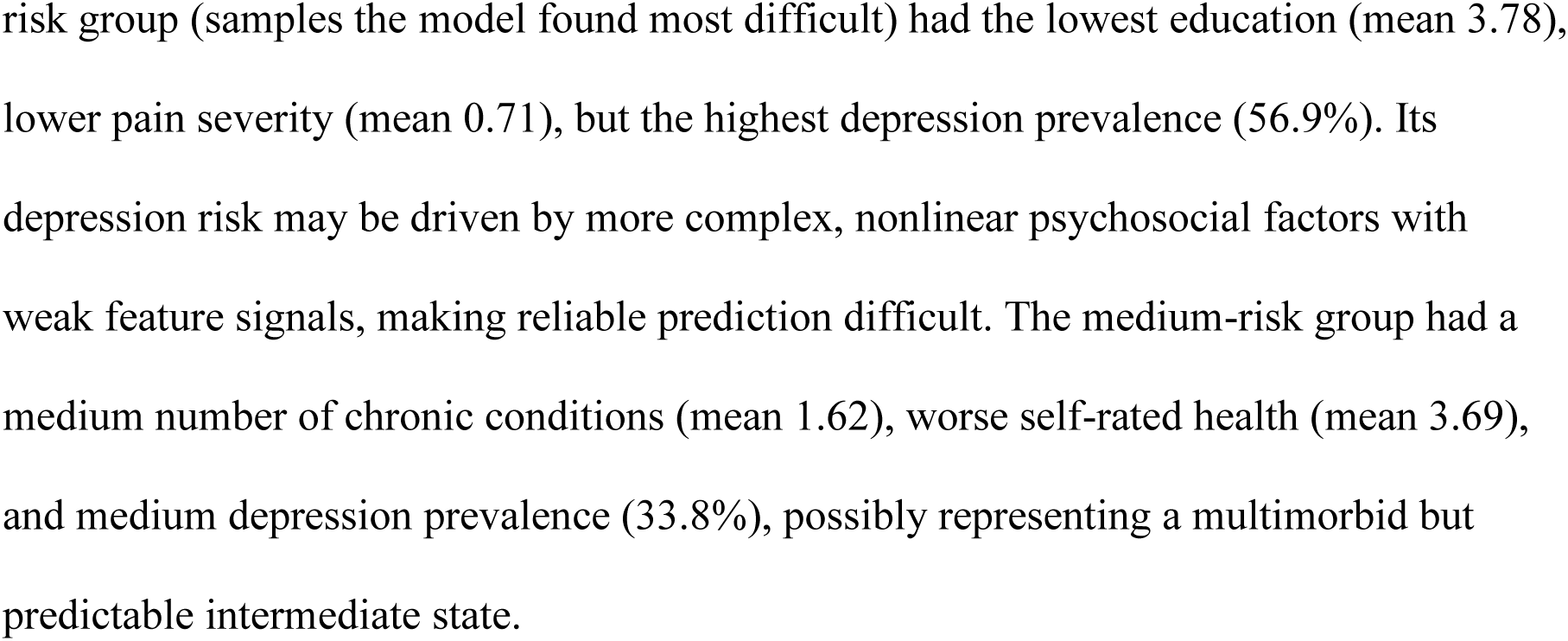
CART risk stratification. Characteristic profiles of low, medium, and high prediction-difficulty groups.

#### CART rule extraction

The CART decision tree produced eight leaf nodes ordered by predicted residual. The leaf-node distribution is provided in Supplementary Table S6.

The rules reveal that self-rated health was the root-node split variable (self_health ≤ 2.5), with education, pain severity, and marital status as subsequent key splits. Participants with lower self-rated health scores (≤2.5) mainly fell into the low-residual region, accounting for about 21% of the sample (905/4,256). Among participants with higher self-rated health (>2.5), those with more education (>7.5 years) also fell into the low-residual region, accounting for about 5% (196/4,256). Those with lower education (≤7.5 years) were further split by pain severity and self-rated health: participants with lower pain (≤1.5) and moderate self-rated health (≤3.5) fell into the medium-residual region, about 19% (824/4,256); participants with lower pain but poorer self-rated health (>3.5) fell into the highest-residual region, about 29% (1,239/4,256). Among participants with higher pain (>1.5), married participants with poorer self-rated health (≤4.5) fell into a medium-high-residual region, whereas those with better self-rated health (>4.5) fell into a lower-residual region. Ordering leaves by predicted residual from low to high, low-residual leaves contained 1,830 samples (43.0%), medium-residual leaves 824 (19.4%), and high-residual leaves 1,602 (37.6%).

#### Multi-parameter and multi-seed stability validation

Multi-parameter stability analysis used five distinct XGBoost hyperparameter configurations (learning_rate, max_depth, n_estimators, subsample, colsample_bytree). Spearman correlations of residuals across the five configurations were all greater than 0.95, indicating high robustness of residuals to hyperparameter variation. Defining the high-risk group as the top 33% of robust residuals, Jaccard similarity of high-risk group sample sets across configurations averaged 0.88, with a minimum of 0.80 and a maximum of 0.94. Multi-seed stability analysis used ten random seeds (0, 1, 2, 3, 4, 21, 42, 84, 100, 123). The mean Jaccard similarity of high-risk group sample sets between any two seeds was 0.88, with a minimum of 0.86 and a maximum of 0.90, and the high-risk group proportion remained stable at approximately 33.0%.

Because the groups were constructed to maximize residual differences, these AUCs are conditional descriptive statistics of the residual-based grouping. The low-risk group achieved LR AUC = 1.000 (95% CI 1.000–1.000, n=298, events=84), the medium-risk group 0.987 (95% CI 0.976–0.995, n=281, events=115), and the high-risk group 0.126 (95% CI 0.088–0.167, n=273, events=156). This extreme divergence is an inherent property of residual-based grouping: the low-risk group consists of samples with the smallest prediction errors, whose depression status is highly separable in feature space; the high-risk group consists of samples that most confuse the model, whose feature-depression relationships are complex and nonlinear and poorly captured by simple logistic regression. From a translational perspective, the low AUC in the high-risk group is not a model failure. It signals that, if these samples were retained, the model would be forced to output probabilities based on uninterpretable black-box guesses, undermining overall interpretability and clinical credibility. Explicitly assigning these samples to a high-uncertainty domain defines a referral pathway: structured human assessment replaces automated scoring, preserving transparency within the interpretable reliable domain. This interpretation should be considered together with the full-sample LR out-of-fold AUC (0.775), within-group mean robust residuals (low 0.162, medium 0.334, high 0.643), and the characteristic profiles. (See Supplementary Figures S4–S7)

**Figure 4.**
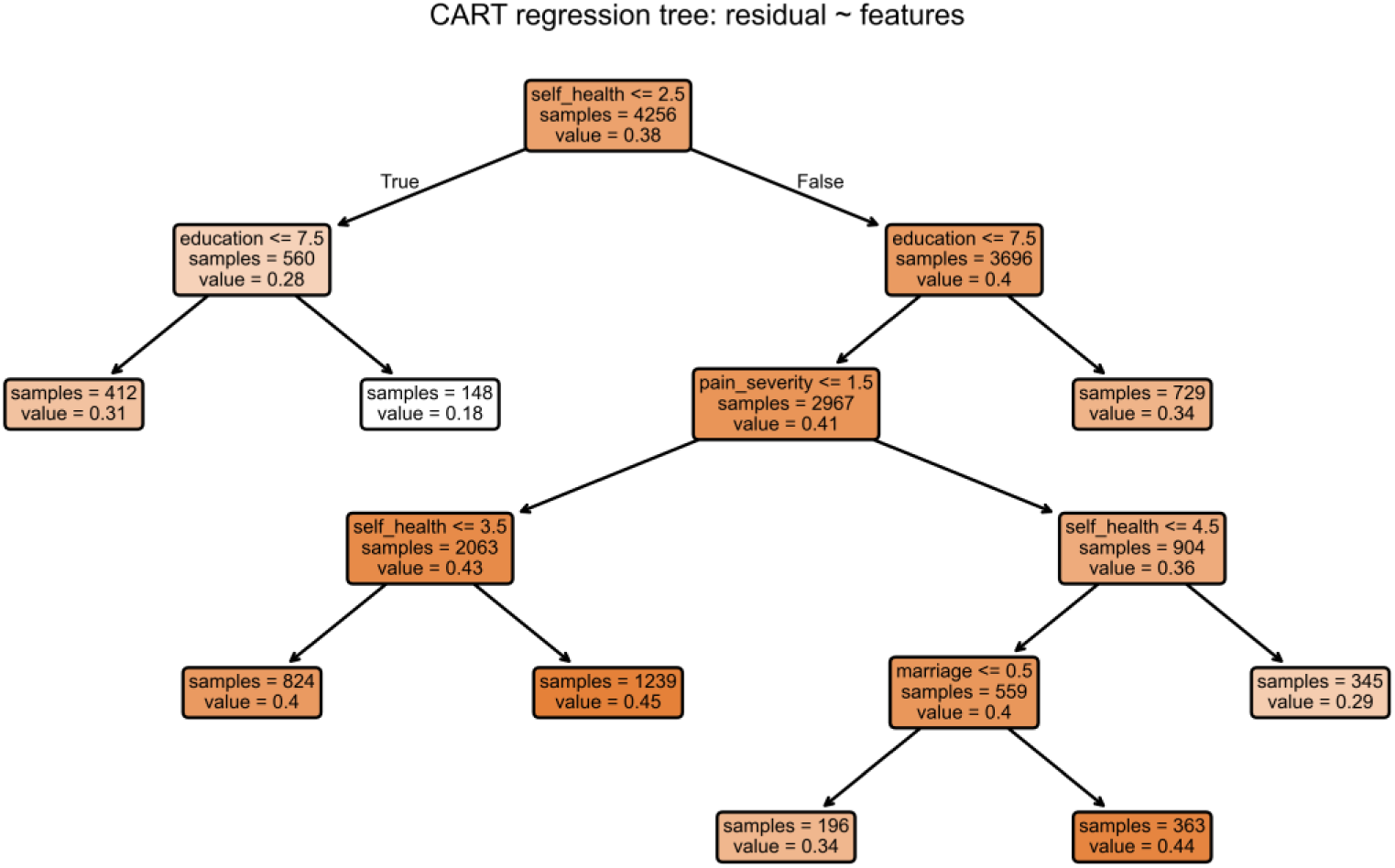
Classification and regression tree rules for robust-residual risk stratification. Split variables are self-rated health, education, pain severity, and marital status.

#### External validation

The harmonized CHARLS-2018 sample comprised 7,964 respondents, with a depression prevalence of 38.2%. XGBoost achieved a full-sample AUC of 0.743. Applying the 2011 CART rules, the low-risk group included 3,998 individuals (50.2%) and had an XGBoost AUC of 0.777; the medium-risk group included 2,024 individuals (25.4%), with a depression prevalence of 30.9% and an XGBoost AUC of 0.611; the high-risk group included 1,942 individuals (24.4%), with a depression prevalence of 57.9% and an XGBoost AUC of 0.609. Group-level AUCs remained stable across multiple XGBoost seeds, indicating a robust operating characteristic.

**Table 6.**
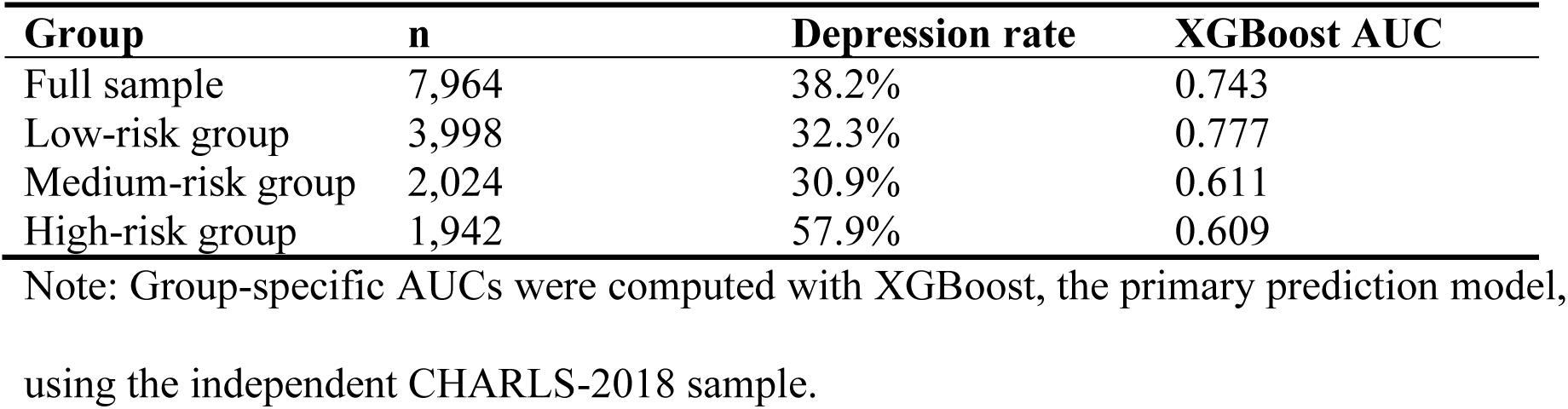
External validation of CART risk groups in CHARLS-2018 (XGBoost).

| Group | n | Depression rate | XGBoost AUC |
| --- | --- | --- | --- |
| Full sample | 7,964 | 38.2% | 0.743 |
| Low-risk group | 3,998 | 32.3% | 0.777 |
| Medium-risk group | 2,024 | 30.9% | 0.611 |
| High-risk group | 1,942 | 57.9% | 0.609 |

Note: Group-specific AUCs were computed with XGBoost, the primary prediction model, using the independent CHARLS-2018 sample.

## Discussion

### A three-step translational chain

#### Step 1: detecting bias: dual distortion created by cumulative selection

CHARLS 2011 provides the empirical base for exposing the dual distortion associated with cumulative inclusion and exclusion criteria in epidemiological inference and machine-learning evaluation. The first distortion affects epidemiological associations. The cancer-depression odds ratio attenuated from 1.78 at L0 to 1.39 at L3, a 21.8% reduction, and lost statistical significance between L1 and L3. Chronic lung disease showed a similar compression pattern. Hypertension and diabetes associations were relatively stable. This differential association compression suggests that selection does not uniformly bias effect estimates; the impact depends on the severity distribution of the exposure and its overlap with exclusion criteria. Collider stratification may be amplified selectively along specific exposure-outcome paths [7].

The second distortion affects machine-learning performance metrics. All five models showed directional increases in AUC with selection strength, but the trends did not survive multiple-comparison correction (family size 20; random forest nominal p=0.044, BH-FDR q=0.148). The rise in AUC therefore appears to reflect mainly a mechanical gain from reduced sample heterogeneity. Selection removed participants with complex, multimorbid, or atypical presentations, leaving a sample with a more compact feature-space distribution and clearer class boundaries, to which AUC as a rank metric is highly sensitive. At the same time, calibration error changed markedly: random forest, XGBoost, LightGBM, and gradient boosting showed significant increases in ECE after BH-FDR correction (q=0.029, 0.001, 0.008, and 0.046), whereas logistic regression ECE decreased. Brier score and mean squared error followed the same pattern: for RF, XGB, LGB, and GB they generally increased with selection, consistent with the ECE deterioration and with mechanical AUC inflation. The combination of a non-significant rise in discrimination and a real degradation in calibration creates a contradictory signal at the metric level. Neither distortion is large enough to trigger alarm on its own. Odds ratios usually move in the same direction, and AUC gains rarely reach obviously implausible levels. Munafo and Smith’s argument about reproducibility applies directly: the compounding effects of subtle design choices are often more damaging than single biases [7]. Formalizing the four-level selection funnel converts that compounding effect from a hidden assumption into a quantifiable empirical object.

A further implication is that selection also constrains the validity domain of model interpretation. Feature weights, SHAP attributions, and CART rules extracted from the selected sample describe relationships that hold within that sample, not necessarily in the source population or in external cohorts.

#### Step 2: mitigating bias: cross-model transfer in the decoupled predictor–selector framework

Once bias is detected, the question becomes how to mitigate it. The model-embedded uncertainty framework offers one path, but its effectiveness depends heavily on the specific pairing of model architecture and uncertainty indicator. Only XGBoost, with its custom out-of-fold quantile residual interval, achieved effective selective prediction: CV coverage approximately 32.4%, test coverage approximately 52.5%, and AUC increased to 0.83. Logistic-regression predictive entropy, random-forest tree-to-tree variance, and LightGBM and gradient boosting 1 − max(probability) all failed to identify an AUC peak better than full coverage. These generic uncertainty indicators lacked the ability to distinguish easy from difficult cases in epidemiological data, and their RC curves were flat, with no informative inflection.

The decoupled predictor–selector framework offered a more robust path. When XGBoost cross-validation residuals were introduced as an external selector, all five models (logistic regression, random forest, XGBoost, LightGBM, and gradient boosting) achieved selective prediction. Cross-validation coverage was highly consistent (approximately 20.0%), CV AUC ranged from 0.8898 to 0.8934, and test AUC ranged from 0.8876 to 0.9008. The models reached cross-architecture consensus on which samples were reliable; they differed only in how those samples were classified. The methodological implication is that information contained in XGBoost cross-validation residuals transfers across model architectures. The quality of uncertainty-indicator design emerges as the critical bottleneck for selective prediction effectiveness.

The robustness of the decoupled framework is further supported by risk-stratification stability. Spearman correlations of residuals across five hyperparameter configurations exceeded 0.95, and multi-seed Jaccard similarity of high-risk groups averaged 0.88 (range 0.86–0.90). Risk stratification can therefore be regarded as a structural property of the data.

#### Step 3: clinical deployment: from black-box rejection to white-box rules

The final step converts methodological findings into a clinically actionable workflow. CART decision-tree splits extracted from XGBoost robust residuals make this conversion possible. Self-rated health was the root-node split variable, dividing the sample into those with poor and better self-rated health. Education, pain severity, and marital status formed subsequent key nodes.

The extracted rules identify three main clinical scenarios. First, participants with self-rated health ≤2.5 mainly fall into the low-residual region, about 21% of the sample. Their depressive symptoms are often linked to clear somatic complaints, feature signals are strong, and model predictions are reliable. Second, participants with self-rated health >2.5 and more than 7.5 years of education also fall into the low-residual region, about 5%. Education here acts as a proxy for cognitive reserve; symptom expression in more educated groups is more easily captured by the available features. Third, participants with self-rated health >2.5, education ≤7.5 years, low pain severity, and poorer self-rated health form the largest high-uncertainty branch, about 29% of the sample; together with the smaller higher-pain high-residual branch, the total high-residual region is about 38%. Depression risk in this group may be driven by more complex psychosocial factors with weak feature signals, and model predictions are less reliable. Viewed through this lens, the CART rule does more than triage patients: it demarcates the boundary between a reliable-interpretation domain and a high-uncertainty domain where model explanations may be distorted by the same selection that compressed the cancer–depression association.

These rules can be embedded directly in primary-care or community-screening settings [28–30]. Clinicians need only four variables (self-rated health score, years of education, pain severity, and marital status) to classify a patient into the model-reliable or model-high-uncertainty domain. Two quantities must be distinguished. The CART-defined low-residual domain (approximately 43%) is a descriptive partition of the whole sample. The decoupled selective-prediction coverage (approximately 20%) selects a high-confidence operating subset from within the tertile-defined low-risk group; 60.1% of that group was selected by the decoupled selector. The former answers ‘for whom is the model suitable?’; the latter decides ‘on whom should automatic scoring be performed.’ For the selected approximately 20% of samples, logistic regression or XGBoost can provide an AUC of approximately 0.89 and precision of approximately 0.86. For the remaining approximately 80% of samples, the model abstains and a human-evaluation workflow is triggered. This triage–predict–stratify workflow turns machine-learning black-box output into white-box decisions based on clinically obtainable variables. Explicitly excluding samples that the current feature set does not adequately learn serves as both a safeguard against misdiagnosis and a requirement for maintaining interpretability: only within a reliable domain do predicted probabilities have clear and stable clinical meaning.

Taken together, bias detection, the decoupled predictor–selector framework, and the CART white-box rules form a logical chain from identifying selection-induced distortion to deploying an interpretable clinical protocol. Each step builds on the previous one, forming a verifiable and reproducible chain. External validity spans temporal, geographic, and domain generalizability [25]; this study interrogates the internal development sample systematically before external validation and exposes the hidden cost of inclusion and exclusion criteria, providing an interpretable answer to ‘generalizable to whom?’

### Domain recommendations

We recommend three reporting practices: (1) replace single-AUC reporting with a conditional tuple (metric, coverage, selection level, calibration trajectory); (2) report OR drift rates across selection levels to assess association robustness; and (3) adopt decoupled selective prediction with an independent, out-of-fold residual selector, accompanied by CART-based risk stratification.

## Limitations

This study has several limitations. The analysis is cross-sectional. Whether the selection-performance relationship persists over time and whether longitudinal attrition introduces additional selection bias cannot be assessed in this design. MICE imputation in L0 may not fully capture uncertainty associated with imputed values. The L0 sample is itself a pseudo-complete population, and its machine-learning metrics are not directly comparable with the complete-case metrics at L1–L3. As a sensitivity analysis, we repeated L0 machine-learning evaluation across five MICE realizations (Supplementary Table S4); test AUC standard deviations were only 0.007–0.009, suggesting that results were insensitive to the choice among imputations, but this does not restore comparability with complete-case levels. Selective-prediction analyses were performed only on the L3 sample. Whether k-nearest-neighbor density selectors or XGB cross-validation residuals generalize to L0 or external populations remains to be validated. The CES-D ≥10 cutoff, although validated in Chinese populations, may miss some clinically relevant depression phenotypes. The binary outcome also precludes analysis of depression severity. CART risk stratification was based only on the L3 sample, so rules may be sample-specific. Rules trained on CHARLS without recalibration may not generalize to HRS or SHARE. The success of XGB cross-validation residuals as an external selector may depend on XGBoost’s specific performance on CHARLS-2011 data. On other datasets, cross-validation residuals from neural networks or other models may transfer better. Despite these limitations, this study provides a methodological template for selection-aware machine-learning evaluation that can be adapted to other observational health datasets.

### Future directions

Several directions follow from this work. Longitudinal extension to an L4 selection level, excluding baseline depression and predicting incident cases, would convert the cross-sectional analysis into a prognostic framework consistent with TRIPOD+AI guidelines [26,27]. Cross-cohort external validation in sister surveys such as HRS and SHARE would test whether the CART rules and the selection–AUC relationships generalize beyond CHARLS or are cohort-specific. Developing predictor–selector decoupling frameworks that separate feature extraction from uncertainty quantification could enable more flexible and safer selective-prediction deployment in clinical settings. Extension to deep neural networks and ensemble methods would reveal whether the coverage variability observed here is amplified or attenuated in more complex architectures. Integrating selective prediction into clinical decision-support systems and packaging model-specific uncertainty profiles as transparent model cards would narrow the gap between methodological research and clinical implementation. Systematic comparison of MICE, joint modeling, and full-information maximum likelihood for missing-data handling in selected samples would clarify how robust the entire pipeline is to imputation strategy. Cross-validation-residual CART risk stratification can be extended directly into clinical decision rules. Model-suitability assessment would then become part of routine clinical practice: every new patient would first be classified by CART rules before receiving a machine-learning prediction.

### Translational significance

The findings converge on a three-tier pathway for embedding selective prediction in chronic-disease management systems [35]. At the entry point (the free annual health examination for residents aged 65 years and older, or routine chronic-disease follow-up visits in primary care), a general practitioner classifies each patient with four routinely collected variables (self-rated health, years of education, pain severity, and marital status) using the CART rules. Patients assigned to the model-reliable domain receive algorithm-assisted depression-risk scoring: at the approximately 20% coverage operating point identified by the decoupled XGBoost residual selector, logistic regression attains an AUC of about 0.89 with precision around 0.86, and these patients remain in primary care for routine follow-up. The remaining approximately 80% of patients, for whom automated predictions are unreliable, are not scored at all; the rule instead flags them for structured human assessment or specialist referral. Coverage therefore functions as a tunable clinical safety boundary.

Viewed through a health-system lens, this triage gate fits the tiered diagnosis and referral structure of Chinese primary care. Embedding the rule in electronic health records and chronic-disease management pathways converts selective prediction from a statistical post-processing step into a routine quality-management checkpoint: algorithmic decisions are issued only under conditions in which the model has demonstrated reliability, and every score carries its conditional reporting tuple (metric, coverage, selection level, calibration trajectory). The resource-allocation implication is direct: in primary-care settings with limited workforce, clinical attention is directed toward the complex subgroup that the model cannot reliably characterize, avoiding unreliable full-coverage prediction.

## Conclusions

Cumulative selection criteria in CHARLS-2011 produced a dual distortion: epidemiological associations (e.g., cancer–depression OR) attenuated, and AUCs rose mechanically as the sample became more homogeneous. Model interpretations derived from the selected sample are similarly conditional, so transparent rules should be reported with their selection level and coverage. A decoupled predictor–selector framework using XGBoost cross-validation residuals achieved an AUC of approximately 0.89 at approximately 20% coverage across five classifiers. CART rules based on self-rated health, education, pain severity, and marital status offer a transparent, four-variable triage tool for primary-care deployment. Conditional performance reporting should become standard practice before such models are used clinically. Embedding such conditional-reporting rules in electronic health records and tiered-care workflows would turn model-uncertainty monitoring into a routine quality-management requirement. In short, the value of this work lies not only in improving individual-level prediction reliability, but also in providing an interpretable and auditable triage node for resource-constrained health systems.

## Abbreviations

ADL: activities of daily living
AUC: area under the receiver operating characteristic curve
CART: classification and regression tree
CES-D: Center for Epidemiologic Studies Depression scale
CHARLS: China Health and Retirement Longitudinal Study
CI: confidence interval
CV: cross-validation
ECE: expected calibration error
FDR: false discovery rate
GB: gradient boosting
IADL: instrumental activities of daily living
LGB: LightGBM
LR: logistic regression
MICE: multiple imputation by chained equations
OR: odds ratio
RC: rejection–coverage
RF: random forest
SMD: standardized mean difference
SMOTE: Synthetic Minority Over-sampling Technique;
STROBE: Strengthening the Reporting of Observational Studies in Epidemiology
TRIPOD: Transparent Reporting of a multivariable prediction model for Individual Prognosis or Diagnosis
XGB: XGBoost.

## Declarations

### Ethics approval and consent to participate

The CHARLS protocol was approved by the Peking University Institutional Review Board (IRB00001052–11015). All participants provided written informed consent.

### Consent for publication

Not applicable

### Availability of data and materials

The data analysed during this study are available from the CHARLS website (https://charls.charlsdata.com/). The analytical code and fitted models are available from the corresponding author upon reasonable request.

### Competing interests

The authors declare no competing interests.

## Funding

This research received no specific grant from any funding agency in the public, commercial, or not-for-profit sectors.

## Authors’ contributions

Zijian Wang conceived and designed the study, performed the data analysis, and drafted the manuscript. Yaqing Liu supervised the study, provided critical feedback, and revised the manuscript. All authors read and approved the final manuscript.

## Data Availability

All data produced are available online at https://charls.charlsdata.com/

## Acknowledgements

We acknowledge the China Health and Retirement Longitudinal Study (CHARLS) team for providing the nationally representative data used in this study. During the preparation of this work, the authors used Kimi (Moonshot AI), Gemini (Google), and ChatGPT (OpenAI) for language editing and text restructuring. The authors reviewed and edited all content and take full responsibility for the integrity of the manuscript.

## Patient and public involvement

No patients or members of the public were involved in the design, conduct, reporting, or interpretation of this study.

## Additional files

Additional file 1: manuscript_Supplementary_Materials (.docx). Supplementary Tables S1–S6 and Supplementary Figures S1–S7. Tables: sample sizes and demographic characteristics by selection level (S1); adjusted odds ratios (95% CI) across selection levels (S2); full machine-learning performance matrix (S3); sensitivity of L0 machine-learning performance to MICE realization (S4); data dictionary and variable codebook (S5); leaf-node distribution of the CART regression tree (S6). Figures: screening-funnel flowchart (S1); missing-data pattern heatmap (S2); heatmap of odds-ratio drift across selection levels (S3); top correlation biases between L0 and L3 (S4); correlation-difference heatmap (S5); distribution of out-of-fold residuals (S6); characteristic profiles of low-, medium-, and high-risk groups (S7).

## Notes

### Competing Interest Statement

The authors have declared no competing interest.

### Author Declarations

The Institutional Review Board of Peking University gave ethical approval for this work (approval number IRB00001052-11015, CHARLS study protocol).

### Summary of Updates

Changes in this revision (v2): 1. The graphical abstract has been removed to comply with the publisher's image-preparation policies. 2. Some references have been added on uncertainty in medical machine learning, prediction-model risk-of-bias assessment, and multiple imputation. 3. An AI-use disclosure has been added to the Acknowledgements. 4. Minor editorial cleanups No results or conclusions have been changed.

