## Supplementary Materials for "Selective prediction as a triage gate for primary-care depression screening: quantifying and mitigating selection bias in CHARLS-2011"

**Supplementary Table S1. Sample sizes and demographic characteristics by selection level (L0 and L3).**

| **Variable** | **Label** | **Type** | **L0 n** | **L3 n** | **L0 mean ± SD** | **L3 mean ± SD** | **SMD** | **SMD magnitude** |
| --- | --- | --- | --- | --- | --- | --- | --- | --- |
| age | Age (years) | continuous | 17705 | 4256 | 59.12 ± 10.02 | 68.62 ± 6.92 | -1.0002974666320312 | Large |
| education | Education (years) | continuous | 17705 | 4256 | 5.31 ± 4.31 | 4.35 ± 4.01 | 0.2267374493904877 | Small |
| cesd10_score | CES-D-10 score | continuous | 17705 | 4256 | 8.44 ± 6.41 | 9.21 ± 6.53 | -0.1198341971533756 | Small |
| chronic_count | Chronic disease count | continuous | 17705 | 4256 | 1.36 ± 1.39 | 1.63 ± 1.47 | -0.189818273825558 | Small |
| self_health | Self-reported health | continuous | 17705 | 4256 | 3.50 ± 1.01 | 3.59 ± 0.99 | -0.0890490506706761 | Negligible |
| pain_severity | Pain severity | continuous | 17705 | 4256 | 0.68 ± 1.09 | 0.72 ± 1.10 | -0.0358816358414673 | Negligible |
| family_support | Family support score | continuous | 17705 | 4256 | 0.00 ± 0.70 | 0.48 ± 0.90 | -0.6378959036128008 | Large |
| log_household_income | Household income (log) | continuous | 17705 | 4256 | 3.66 ± 4.26 | 3.16 ± 4.07 | 0.1183193667667901 | Small |
| log_individual_income | Individual income (log) | continuous | 17705 | 4256 | 0.59 ± 2.22 | 0.23 ± 1.38 | 0.1719356331158477 | Small |
| gender | Female, n (%) | binary | 17705 | 4256 | 8,471 (47.8%) | 2,294 (53.9%) | -0.1213438687089265 | Small |
| marriage | Married/partnered, n (%) | binary | 17705 | 4256 | 15,417 (87.1%) | 2,885 (67.8%) | 0.4742570640814865 | Moderate |
| residence | Urban residence, n (%) | binary | 17705 | 4256 | 4,303 (24.3%) | 1,039 (24.4%) | -0.0025329471969063 | Negligible |
| has_insurance | Has medical insurance, n (%) | binary | 17705 | 4256 | 16,447 (92.9%) | 3,950 (92.8%) | 0.0032805240217299 | Negligible |
| depression | Depression (CES-D >= 10), n (%) | binary | 17705 | 4256 | 6,534 (36.9%) | 1,772 (41.6%) | -0.0969807681085322 | Negligible |
| adl_any | Any ADL limitation, n (%) | binary | 17705 | 4256 | 2,589 (14.6%) | 776 (18.2%) | -0.0975464424781525 | Negligible |
| live_with_child | Living with child, n (%) | binary | 17705 | 3816 | 1,933 (10.9%) | 699 (18.3%) | -0.2106156453308715 | Small |
| hypertension | Hypertension, n (%) | binary | 17705 | 4256 | 4,284 (24.2%) | 1,364 (32.0%) | -0.1753217023417526 | Small |
| diabetes_or_highbs | Diabetes/high blood sugar, n (%) | binary | 17705 | 4256 | 993 (5.6%) | 291 (6.8%) | -0.050883991023946 | Negligible |
| chronic_cancer | Cancer, n (%) | binary | 17705 | 4256 | 180 (1.0%) | 41 (1.0%) | 0.0053852011798532 | Negligible |
| chronic_lung_disease | Chronic lung disease, n (%) | binary | 17705 | 4256 | 1,781 (10.1%) | 606 (14.2%) | -0.1281923499391622 | Small |

**Supplementary Table S2. Adjusted odds ratios (95% CI) and L0→L3 drift rates for all chronic-disease exposures across selection levels.**

| **Exposure** | **L0 OR (95% CI)** | **L1 OR (95% CI)** | **L2 OR (95% CI)** | **L3 OR (95% CI)** | **L0→L3 drift (%)** | **L0 n** | **L1 n** | **L2 n** | **L3 n** |
| --- | --- | --- | --- | --- | --- | --- | --- | --- | --- |
| Cancer | 1.781 [1.317–2.410] | 1.315 [0.799–2.164] | 1.371 [0.816–2.301] | 1.393 [0.737–2.632] | -21.8% | 17705 | 6920 | 6594 | 4256 |
| Chronic disease count | 1.450 [1.416–1.486] | 1.416 [1.367–1.468] | 1.412 [1.361–1.464] | 1.422 [1.358–1.489] | -1.9% | 17705 | 6920 | 6594 | 4256 |
| Chronic lung disease | 1.998 [1.804–2.214] | 1.749 [1.519–2.013] | 1.754 [1.518–2.027] | 1.883 [1.575–2.251] | -5.8% | 17705 | 6920 | 6594 | 4256 |
| Diabetes/high blood sugar | 1.414 [1.237–1.616] | 1.424 [1.176–1.723] | 1.395 [1.147–1.696] | 1.433 [1.119–1.834] | 1.3% | 17705 | 6920 | 6594 | 4256 |
| Hypertension | 1.255 [1.165–1.350] | 1.195 [1.075–1.327] | 1.180 [1.059–1.315] | 1.214 [1.062–1.388] | -3.3% | 17705 | 6920 | 6594 | 4256 |

**Supplementary Table S3. Full machine-learning performance matrix (mean ± SD across seven random seeds) by model and selection level.**

| **Level** | **Model** | **Test AUC** | **Test Brier** | **Test ECE** | **Test MSE** |
| --- | --- | --- | --- | --- | --- |
| L0 | Gradient Boosting | 0.776 ± 0.005 | 0.183 ± 0.002 | 0.045 ± 0.007 | 0.183 ± 0.002 |
| L0 | LightGBM | 0.776 ± 0.005 | 0.183 ± 0.002 | 0.045 ± 0.007 | 0.183 ± 0.002 |
| L0 | Logistic Regression | 0.776 ± 0.006 | 0.192 ± 0.003 | 0.104 ± 0.005 | 0.192 ± 0.003 |
| L0 | Random Forest | 0.774 ± 0.006 | 0.185 ± 0.002 | 0.062 ± 0.003 | 0.185 ± 0.002 |
| L0 | XGBoost | 0.775 ± 0.005 | 0.183 ± 0.002 | 0.048 ± 0.007 | 0.183 ± 0.002 |
| L1 | Gradient Boosting | 0.770 ± 0.021 | 0.192 ± 0.007 | 0.050 ± 0.009 | 0.192 ± 0.007 |
| L1 | LightGBM | 0.769 ± 0.021 | 0.192 ± 0.007 | 0.045 ± 0.006 | 0.192 ± 0.007 |
| L1 | Logistic Regression | 0.771 ± 0.020 | 0.194 ± 0.007 | 0.067 ± 0.008 | 0.194 ± 0.007 |
| L1 | Random Forest | 0.766 ± 0.021 | 0.195 ± 0.008 | 0.066 ± 0.010 | 0.195 ± 0.008 |
| L1 | XGBoost | 0.769 ± 0.021 | 0.192 ± 0.007 | 0.049 ± 0.008 | 0.192 ± 0.007 |
| L2 | Gradient Boosting | 0.773 ± 0.010 | 0.190 ± 0.003 | 0.053 ± 0.010 | 0.190 ± 0.003 |
| L2 | LightGBM | 0.773 ± 0.010 | 0.190 ± 0.003 | 0.053 ± 0.008 | 0.190 ± 0.003 |
| L2 | Logistic Regression | 0.776 ± 0.010 | 0.192 ± 0.003 | 0.070 ± 0.007 | 0.192 ± 0.003 |
| L2 | Random Forest | 0.773 ± 0.009 | 0.194 ± 0.003 | 0.074 ± 0.011 | 0.194 ± 0.003 |
| L2 | XGBoost | 0.773 ± 0.010 | 0.190 ± 0.003 | 0.055 ± 0.006 | 0.190 ± 0.003 |
| L3 | Gradient Boosting | 0.783 ± 0.023 | 0.187 ± 0.009 | 0.061 ± 0.008 | 0.187 ± 0.009 |
| L3 | LightGBM | 0.783 ± 0.022 | 0.187 ± 0.009 | 0.064 ± 0.008 | 0.187 ± 0.009 |
| L3 | Logistic Regression | 0.785 ± 0.024 | 0.189 ± 0.009 | 0.079 ± 0.012 | 0.189 ± 0.009 |
| L3 | Random Forest | 0.782 ± 0.025 | 0.191 ± 0.008 | 0.083 ± 0.022 | 0.191 ± 0.008 |
| L3 | XGBoost | 0.783 ± 0.022 | 0.187 ± 0.009 | 0.061 ± 0.011 | 0.187 ± 0.009 |

**Supplementary Table S4. Sensitivity of L0 machine-learning performance to the choice of MICE realization.**

| **Model** | **MICE realization** | **AUC** | **Brier** | **ECE** | **MSE** |
| --- | --- | --- | --- | --- | --- |
| GB | 5 | 0.777 ± 0.007 | 0.182 ± 0.003 | 0.046 ± 0.003 | 0.182 ± 0.003 |
| LGB | 5 | 0.777 ± 0.007 | 0.182 ± 0.003 | 0.045 ± 0.002 | 0.182 ± 0.003 |
| LR | 5 | 0.777 ± 0.009 | 0.191 ± 0.004 | 0.104 ± 0.004 | 0.191 ± 0.004 |
| RF | 5 | 0.775 ± 0.008 | 0.185 ± 0.003 | 0.060 ± 0.002 | 0.185 ± 0.003 |
| XGB | 5 | 0.776 ± 0.007 | 0.182 ± 0.003 | 0.047 ± 0.004 | 0.182 ± 0.003 |

**Supplementary Table S5. Data dictionary and variable codebook for the CHARLS-2011 analysis dataset.**

| **Field** | **Label** | **Type** | **Missing rate** | **Non-missing n** | **Unique values** |
| --- | --- | --- | --- | --- | --- |
| age | Age (2011 - birth year) | float64 | 0.00% | 17705 | 55 |
| gender | Sex, 0=female, 1=male | float64 | 0.00% | 17705 | 2 |
| education | Years of education (continuous) | float64 | 0.00% | 17705 | 9 |
| marriage | Marital status, 0=unmarried/divorced/widowed, 1=married | int64 | 0.00% | 17705 | 2 |
| ID | Individual ID | str | 0.00% | 17705 | 17705 |
| householdID | Household ID | str | 0.00% | 17705 | 10251 |
| communityID | Community ID | str | 0.00% | 17705 | 450 |
| residence | Urban/rural residence, 0=rural, 1=urban | float64 | 42.27% | 10221 | 2 |
| live_with_child | Living with children, 0/1 | float64 | 26.61% | 12994 | 2 |
| money_amount | Money received from children (raw value) | float64 | 82.80% | 3045 | 212 |
| log_money_amount | Money received from children, log1p-transformed | float64 | 42.83% | 10122 | 210 |
| got_money | Received financial support from children, 0/1 | float64 | 42.83% | 10122 | 2 |
| goods_amount | Goods received from children (raw value) | float64 | 92.38% | 1349 | 128 |
| log_goods_amount | Goods received from children, log1p-transformed | float64 | 42.83% | 10122 | 128 |
| got_goods | Received in-kind support from children, 0/1 | float64 | 42.83% | 10122 | 2 |
| got_transfer | Received any transfers (money + goods), 0/1 | float64 | 42.83% | 10122 | 2 |
| depression | Depression label, 1=depressed, 0=not depressed | float64 | 10.38% | 15867 | 2 |
| cesd10_score | CESD-10 continuous score (sum of non-missing items) | float64 | 10.38% | 15867 | 31 |
| cesd_bothered | CESD-10: bothered by things in the past week (dc009) | float64 | 10.09% | 15919 | 4 |
| cesd_concentrate | CESD-10: trouble concentrating in the past week (dc010) | float64 | 10.63% | 15823 | 4 |
| cesd_depressed | CESD-10: felt depressed in the past week (dc011) | float64 | 10.43% | 15858 | 4 |
| cesd_effort | CESD-10: everything was an effort in the past week (dc012) | float64 | 10.26% | 15888 | 4 |
| cesd_hopeful | CESD-10: felt hopeful about the future in the past week (dc013, reverse-scored) | float64 | 11.73% | 15629 | 4 |
| cesd_fearful | CESD-10: felt fearful in the past week (dc014) | float64 | 9.80% | 15970 | 4 |
| cesd_sleep | CESD-10: restless sleep in the past week (dc015) | float64 | 9.69% | 15989 | 4 |
| cesd_happy | CESD-10: felt happy in the past week (dc016, reverse-scored) | float64 | 9.94% | 15946 | 4 |
| cesd_lonely | CESD-10: felt lonely in the past week (dc017) | float64 | 10.07% | 15922 | 4 |
| cesd_getgoing | CESD-10: could not get going in the past week (dc018) | float64 | 10.29% | 15884 | 4 |
| chronic_count | Number of chronic diseases (14 conditions) | float64 | 0.00% | 17705 | 11 |
| hypertension | Hypertension, 0/1 | float64 | 0.00% | 17705 | 2 |
| diabetes_or_highbs | Diabetes or high blood sugar, 0/1 (da007_3) | float64 | 0.00% | 17705 | 2 |
| chronic_cancer | Cancer, 0/1 | float64 | 0.00% | 17705 | 2 |
| chronic_lung_disease | Chronic lung disease, 0/1 (da007_5) | float64 | 0.00% | 17705 | 2 |
| self_health | Self-rated health, 1=best, 5=worst; da001 preferred, da002 used when missing | float64 | 0.00% | 17705 | 5 |
| adl_any | ADL/IADL limitation (>=3 items), 0/1 | float64 | 0.00% | 17705 | 2 |
| pain_severity | Pain severity, 0=none, 1=mild, 2=moderate, 3=severe | float64 | 0.00% | 17705 | 4 |
| has_insurance | Has health insurance, 0/1 | float64 | 0.00% | 17705 | 2 |
| insurance_type | Insurance type code (5=government, 1=employee, 3=rural cooperative, 2=resident, etc.) | float64 | 8.01% | 16287 | 9 |
| individual_income | Individual total income (truncated) | float64 | 0.00% | 17705 | 86 |
| log_individual_income | Individual total income, log1p-transformed | float64 | 0.00% | 17705 | 88 |
| household_income | Household total income (truncated, incl. agricultural output and self-employment net income) | float64 | 0.00% | 17705 | 231 |
| log_household_income | Household total income, log1p-transformed | float64 | 0.00% | 17705 | 231 |
| family_support | Family support composite score (standardized, equal-weighted sum) | float64 | 0.00% | 17705 | 409 |
| age_group | Age group (<60 / 60-74 / >=75) | category | 0.00% | 17705 | 3 |
| edu_group | Education group (primary school or below / middle school / high school or above) | category | 0.00% | 17705 | 3 |
| support_group | Family support tertile (low / medium / high) | category | 0.00% | 17705 | 3 |

**Supplementary Table S6. Leaf-node distribution of the CART regression tree fitted to robust residuals (n = 4,256).**

| **Leaf ID** | **Dominant risk group** | **n** | **Mean robust residual** | **Low (%)** | **Medium (%)** | **High (%)** |
| --- | --- | --- | --- | --- | --- | --- |
| 10 | Low | 148 | 0.18 | 82.4% | 8.8% | 8.8% |
| 8 | Low | 345 | 0.288 | 66.7% | 14.8% | 18.6% |
| 9 | Low | 412 | 0.311 | 61.7% | 15.8% | 22.6% |
| 13 | Low | 196 | 0.335 | 51.5% | 24.5% | 24.0% |
| 4 | Low | 729 | 0.342 | 47.1% | 26.7% | 26.2% |
| 11 | Medium | 824 | 0.396 | 24.0% | 45.4% | 30.6% |
| 14 | High | 363 | 0.442 | 19.6% | 36.1% | 44.4% |
| 12 | High | 1239 | 0.451 | 8.0% | 43.7% | 48.3% |

**Supplementary Figure S1. Screening funnel flowchart showing sample sizes and exclusion criteria at each level (L0–L3).**

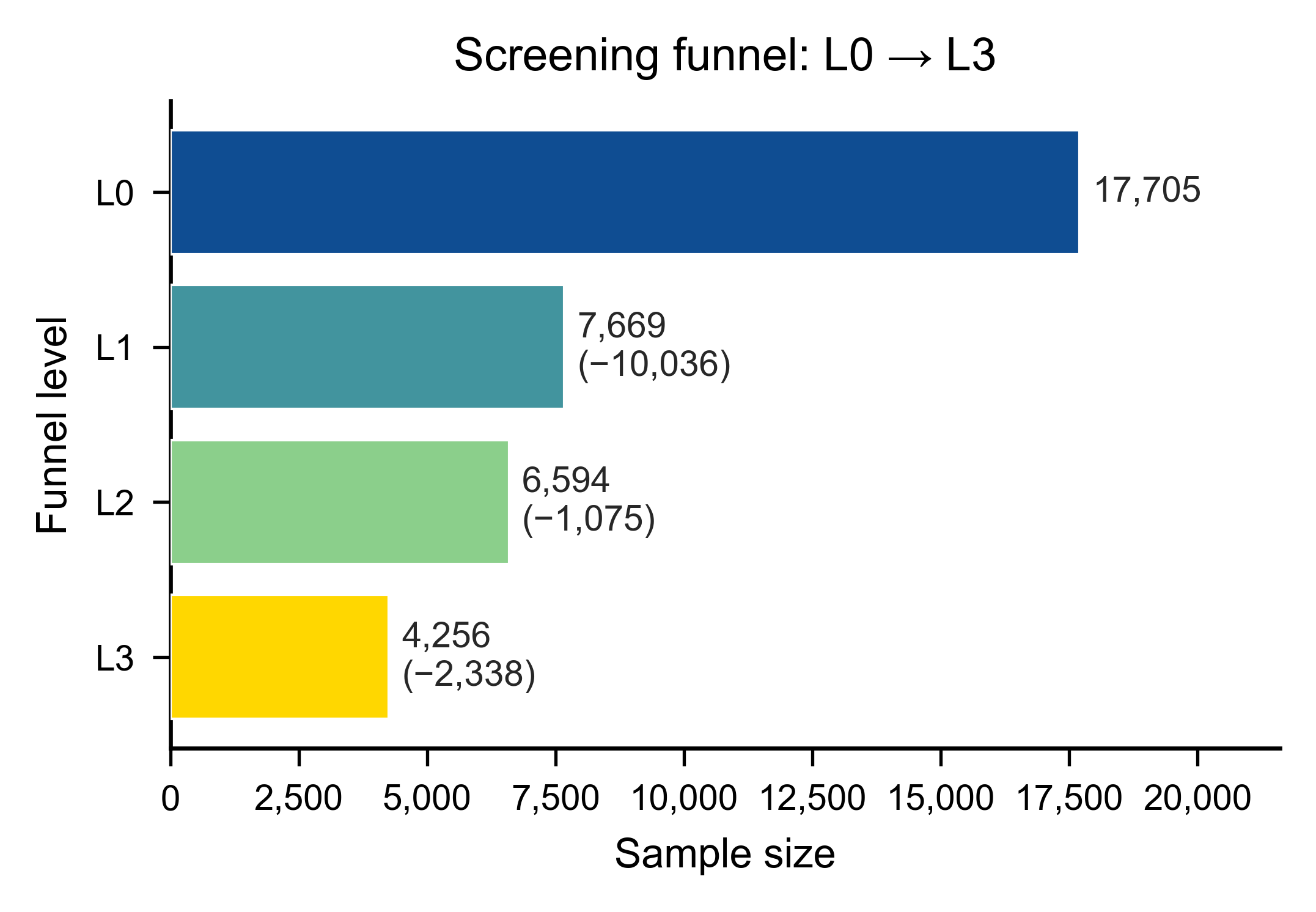

**Supplementary Figure S2. Missing-data pattern heatmap for variables used in the L0–L3 funnel.**

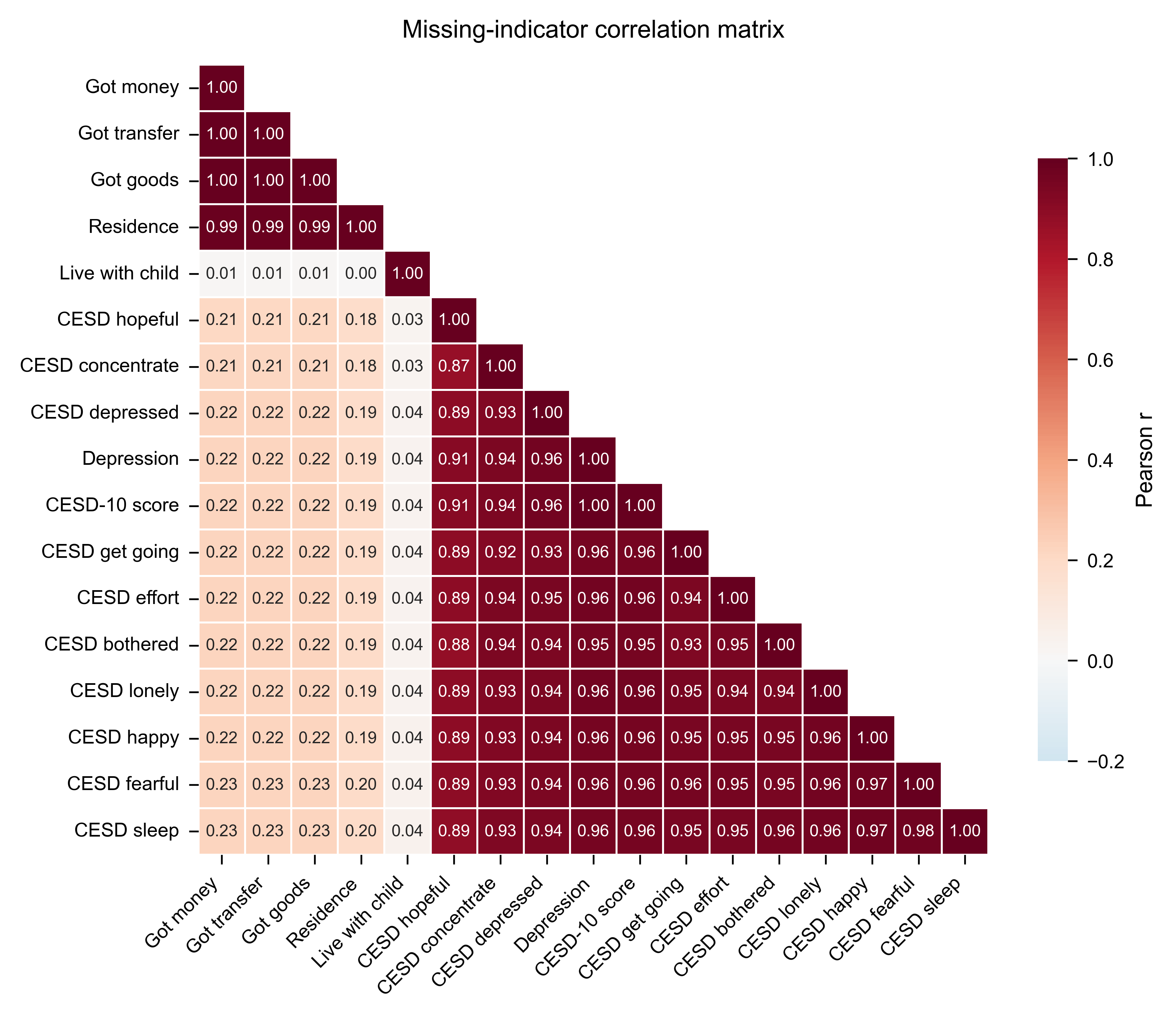

**Supplementary Figure S3. Heatmap of odds-ratio drift across selection levels for all chronic-disease exposures.**

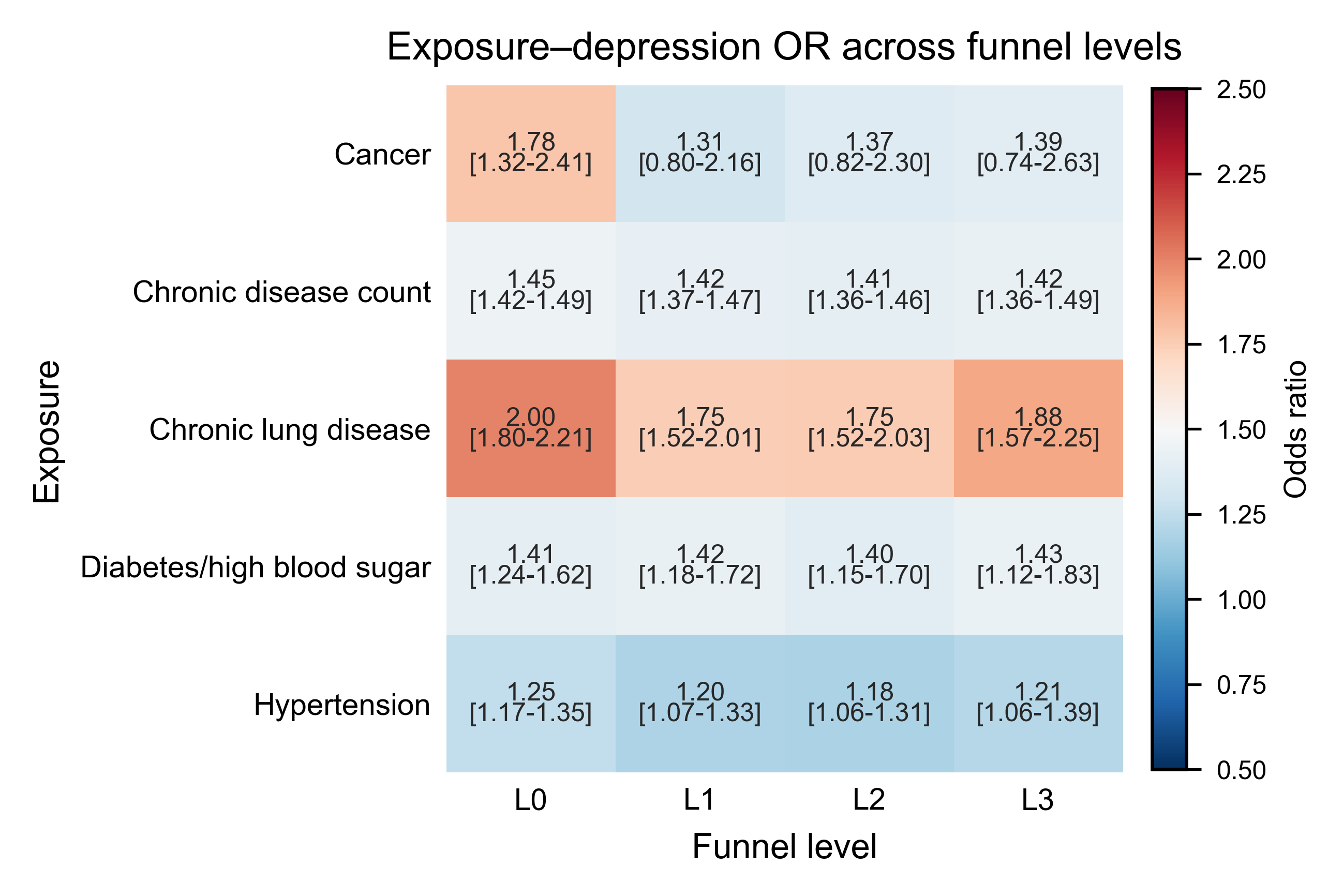

**Supplementary Figure S4. Top 6 correlation biases between L0 (MICE-imputed) and L3 (most selective) samples.**

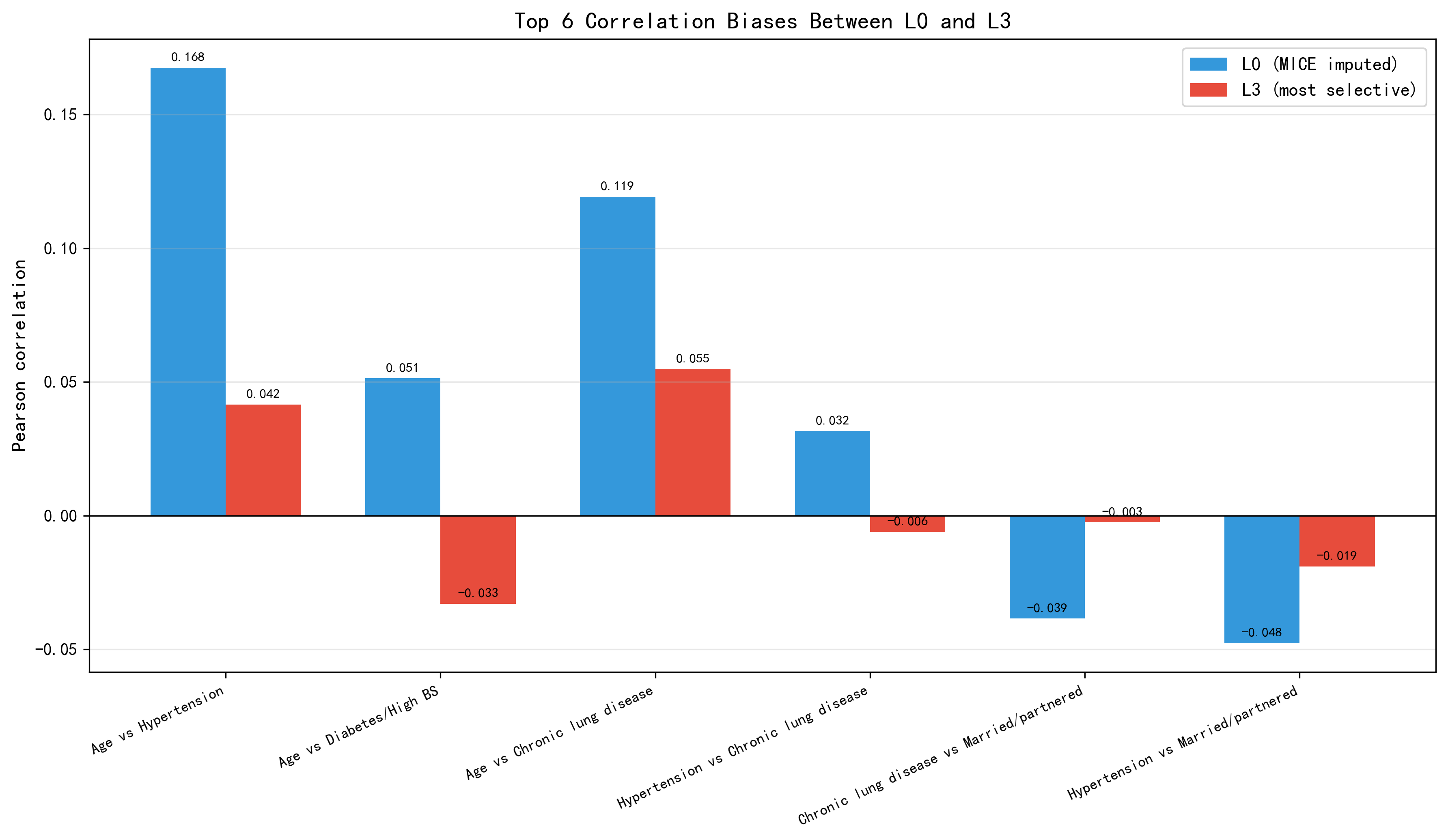

**Supplementary Figure S5. Correlation-difference heatmap (L3 minus L0) for selected variables.**

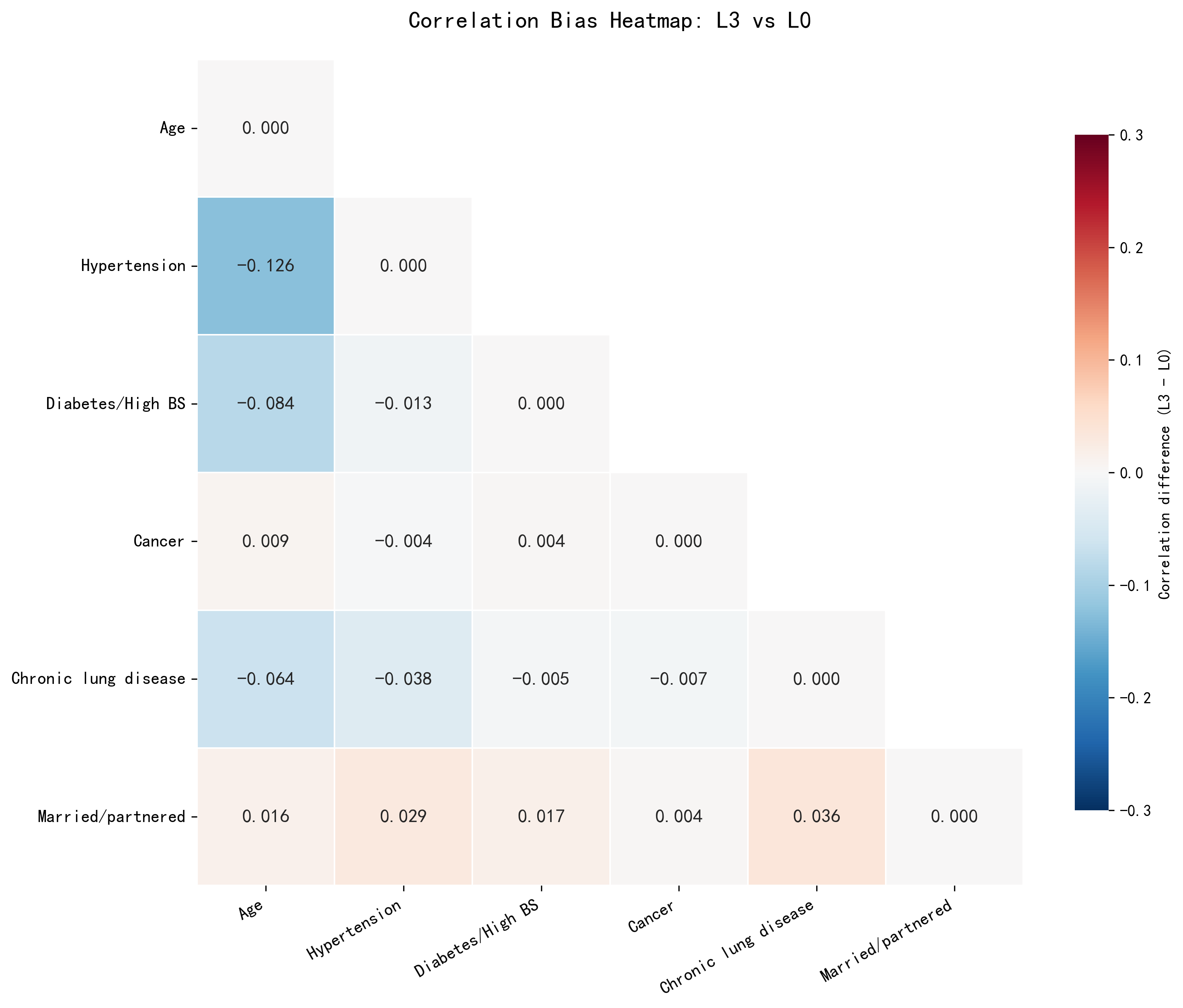

**Supplementary Figure S6. Distribution of out-of-fold residuals across five XGBoost hyperparameter configurations.**

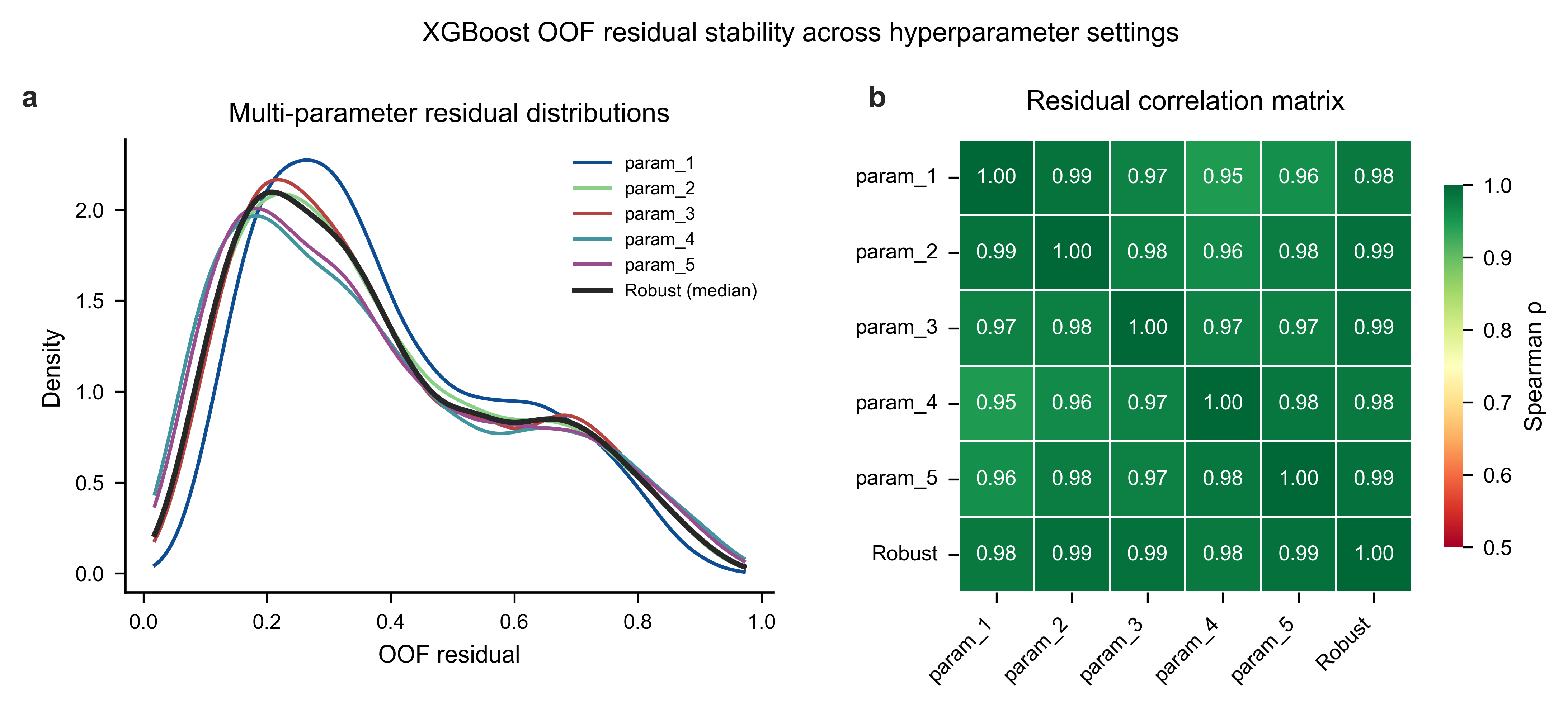

**Supplementary Figure S7. Characteristic profiles of low-, medium-, and high-risk groups defined by robust residuals.**

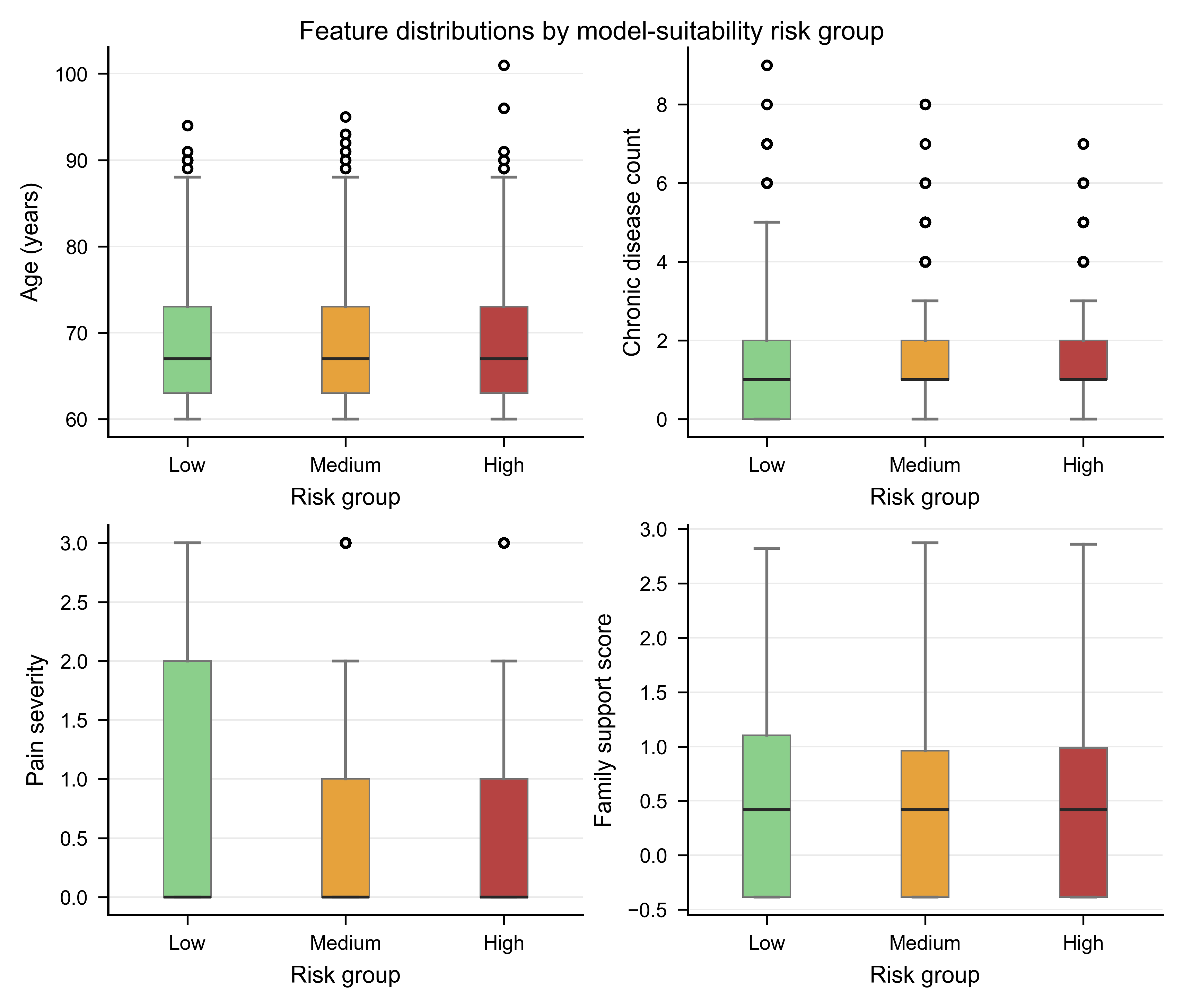
